# The blood metabolome of cognitive function and brain health in middle-aged adults – influences of genes, gut microbiome, and exposome

**DOI:** 10.1101/2024.12.16.24317793

**Authors:** Shahzad Ahmad, Tong Wu, Matthias Arnold, Thomas Hankemeier, Mohsen Ghanbari, Gennady Roshchupkin, André G. Uitterlinden, Julia Neitzel, Robert Kraaij, Cornelia M. Van Duijn, M. Arfan Ikram, Rima Kaddurah-Daouk, Gabi Kastenmüller, the Alzheimer’s Disease Metabolomics Consortium

## Abstract

Increasing evidence suggests the involvement of metabolic alterations in neurological disorders, including Alzheimer’s disease (AD), and highlights the significance of the peripheral metabolome, influenced by genetic factors and modifiable environmental exposures, for brain health. In this study, we examined 1,387 metabolites in plasma samples from 1,082 dementia-free middle-aged participants of the population-based Rotterdam Study. We assessed the relation of metabolites with general cognition (G-factor) and magnetic resonance imaging (MRI) markers using linear regression and estimated the variance of these metabolites explained by genes, gut microbiome, lifestyle factors, common clinical comorbidities, and medication using gradient boosting decision tree analysis. Twenty-one metabolites and one metabolite were significantly associated with total brain volume and total white matter lesions, respectively. Fourteen metabolites showed significant associations with G-factor, with ergothioneine exhibiting the largest effect (adjusted mean difference = 0.122, *P* = 4.65x10^-7^). Associations for nine of the 14 metabolites were replicated in an independent, older cohort. The metabolite signature of incident AD in the replication cohort resembled that of cognition in the discovery cohort, emphasizing the potential relevance of the identified metabolites to disease pathogenesis. Lifestyle, clinical variables, and medication were most important in determining these metabolites’ blood levels, with lifestyle, explaining up to 28.6% of the variance. Smoking was associated with ten metabolites linked to G-factor, while diabetes and antidiabetic medication were associated with 13 metabolites linked to MRI markers, including N-lactoyltyrosine. Antacid medication strongly affected ergothioneine levels. Mediation analysis revealed that lower ergothioneine levels may partially mediate negative effects of antacids on cognition (31.5%). Gut microbial factors were more important for the blood levels of metabolites that were more strongly associated with cognition and incident AD in the older replication cohort (beta-cryptoxanthin, imidazole propionate), suggesting they may be involved later in the disease process. The detailed results on how multiple modifiable factors affect blood levels of cognition- and brain imaging-related metabolites in dementia-free participants may help identify new AD prevention strategies.

## INTRODUCTION

Growing evidence implicates alterations of the peripheral metabolism in brain-related diseases including Alzheimer’s disease (AD) (1-3). Multiple studies have linked the blood levels of various metabolites, including amino acids (leucine, isoleucine, glutamine, valine), lipids (fatty acids, acylcarnitines, phosphatidylcholines, sphingomyelins, bile acids), and lipoproteins, with AD-related phenotypes such as cognition and brain imaging phenotypes, or the conversion from a cognitively normal state to AD (4-15).

Highlighting the long pre-symptomatic phase of AD, structural and functional changes in various brain regions and alterations in neuropsychological markers emerge many years before the manifestation of the disease (16-19). In particular, brain atrophy (cortical thinning, reduction in hippocampal volume), and the presence of white matter hyperintensities as detected through magnetic resonance imaging (MRI) serve as distinctive neurodegenerative and vascular imaging markers associated with AD (20-22). Studying the metabolic changes linked with these features of AD early, before the onset of any symptoms might help to identify metabolites that are important in the etiology of the disease.

Blood metabolomes reflect metabolic states as resulting from the interplay between genetics, gut microbiome, and exposome (23, 24). In recent studies, the gut microbiome and the exposome, which includes lifestyle factors, medication use, and further environmental exposures, emerged as important contributing factors to influence metabolite levels in addition to genetics (25-28). For example, by building a reference map of potential determinants of the human serum metabolome in an Israeli cohort (25), diet and gut microbiome were shown to play a crucial role in defining the metabolic repertoire within the systemic circulation. Metabolites co-metabolized by gut microbiota may not only serve as nutrients for microbial flora and the host, but also as signaling molecules to regulate host molecular pathways (29).

As the gut microbiome and many factors of the exposome are modifiable in nature, their management provides an opportunity to stabilize metabolism and counteract disease-related metabolic alterations. However, to define targeted interventions and prevention strategies, a profound understanding is required of (i) the metabolites associated with early alterations in brain structure and cognitive function related to AD, and (ii) the extent to which these metabolite levels are influenced (and thus potentially modifiable) by the gut microbiome and the exposome in pre-symptomatic participants.

In this study, we first investigated the association of metabolites with general cognition and magnetic resonance imaging markers of AD in a dementia-free sample of the prospective population-based Rotterdam Study (RS). In a second step, we evaluated the contribution of genetic variation, gut microbiome, various lifestyle factors, medication use, and clinical features to define levels of identified metabolites in circulation and investigated the interplay of these factors in detail for selected examples, exploring their potential as targets for intervention and prevention.

## RESULTS

For this study, the levels of 1,387 metabolites were determined in plasma samples from 1,082 participants of the RSIII-2 cohort (i.e., the third cohort at second follow-up) of the Rotterdam Study using a non-targeted metabolomics approach (Metabolon HD4). To elucidate the connection of the peripheral metabolome with brain health and function, we first investigated the association of general cognition and MRI markers with the levels of 991 frequent metabolites in 1,068 participants (after data preprocessing) (see **Methods**; **Supplementary Figure 1**). For cognition, we replicated our findings in 874 participants of the older RSI-4 cohort, an independent sample (first cohort) of the Rotterdam Study, for which metabolomics data from the same platform were available. The characteristics of participants from the RSIII-2 (N = 1,068) and RSI-4 (N = 874) cohorts are summarized in **Table 1**. Participants of both cohorts were dementia-free at the time of blood sampling.

**Table 1:** Population characteristics.

| Study characteristics | Rotterdam Study III<br>(N = 1068) | Rotterdam Study I<br>(N = 874) |
| --- | --- | --- |
| Age in years (mean, SD) | 62.54 (5.91) | 76.25 (4.78) |
| Sex (Female %) | 598 (56) | 506 (57.89) |
| Body mass index in kg/m<sup>2</sup> (mean, SD) | 27.35 (4.30) | 27.54 (4.23) |
| Smoking status (%) |  |  |
| Never | 350 (32.56) | 250 (28.60) |
| Former | 574 (54.42) | 531 (60.75) |
| Current | 140 (13.02) | 93 (10.64) |
| Educational attainment (%) |  |  |
| Primary education | 422 (39.62) | 519 (59.93) |
| Further education | 308 (28.92) | 257 (29.67) |
| Higher education | 335 (31.45) | 90 (10.39) |
| Hypertension (%) | 497 (46.66) | 506 (57.89) |
| Total cholesterol (mmol/L) [Mean, SD] | 5.578 (1.12) | 5.56 (0.95) |
| HDL cholesterol (mmol/L) [Mean, SD] | 1.507 (0.45) | 1.44 (0.39) |
| Use of lipid lowering drugs (%) | 286 (12) | 186 (21.16) |
| Diabetes (%) | 65 (6.09) | 62 (7.11) |

To explore the potential of modifying the cognition- and MRI markers-associated metabolites through interventions, we assessed how much metabolite levels were influenced by modifiable versus unmodifiable features as a second step. For this purpose, we estimated the portion of metabolite variance that is explained by the variation in the participants’ gut microbiome, lifestyle, clinical factors, medication, and genetics for all metabolites, and additionally investigated the pairwise association between each factor and metabolite. Details of the performed analyses are provided in the **Methods** section. A general overview over the steps is given in **Supplementary Figure 2**.

### General cognition and brain MRI markers are associated with distinct blood metabolites in a population-based cohort

Analyzing the relationship of 991 metabolites with general cognition, we observed significant associations (false discovery rate (FDR) < 0.05) of 14 metabolites with cognition while adjusting for age, sex, body mass index (BMI), and lipid-lowering medication use (model 1) (**Figure 1**, **Table 2, Supplementary Table 1)**. Higher levels of ergothioneine (adjusted mean difference = 0.122, *P* = 4.65x10^-7^), uridine (0.093, *P* = 1.0x10^-4^), 2-deoxyuridine (0.083, *P* = 5.48x10^-4^), and two chemically uncharacterized metabolites (X – 11849, X – 11847) were associated with better cognition. Moreover, lower levels of seven sulfated xenobiotic metabolites (4-vinylguaiacol sulfate, o-cresol sulfate, 3-acetylphenol sulfate, 3-hydroxy-2-methylpyridine sulfate, 2-naphthol sulfate, 4-vinylcatechol sulfate, 3-methylcatechol sulfate) and two uncharacterized metabolites (X – 25420, X – 24418) showed association with better general cognition. These metabolites fall into three main sets of correlated metabolites (**Supplementary Figure 3**), namely (i) sulfated xenobiotics (0.47 < *r* < 0.79), (ii) the nucleotides 2’-deoxyuridine and uridine (*r* = 0.37) and ergothioneine (*r* = 0.27), and (iii) the uncharacterized metabolites X – 11849, X – 11847 (r = 0.86). After adjustment for education (model 2) (**Supplementary Table 1**), the associations for 12 out of the 14 metabolites remained significant (FDR < 0.05).

**Figure 1:**
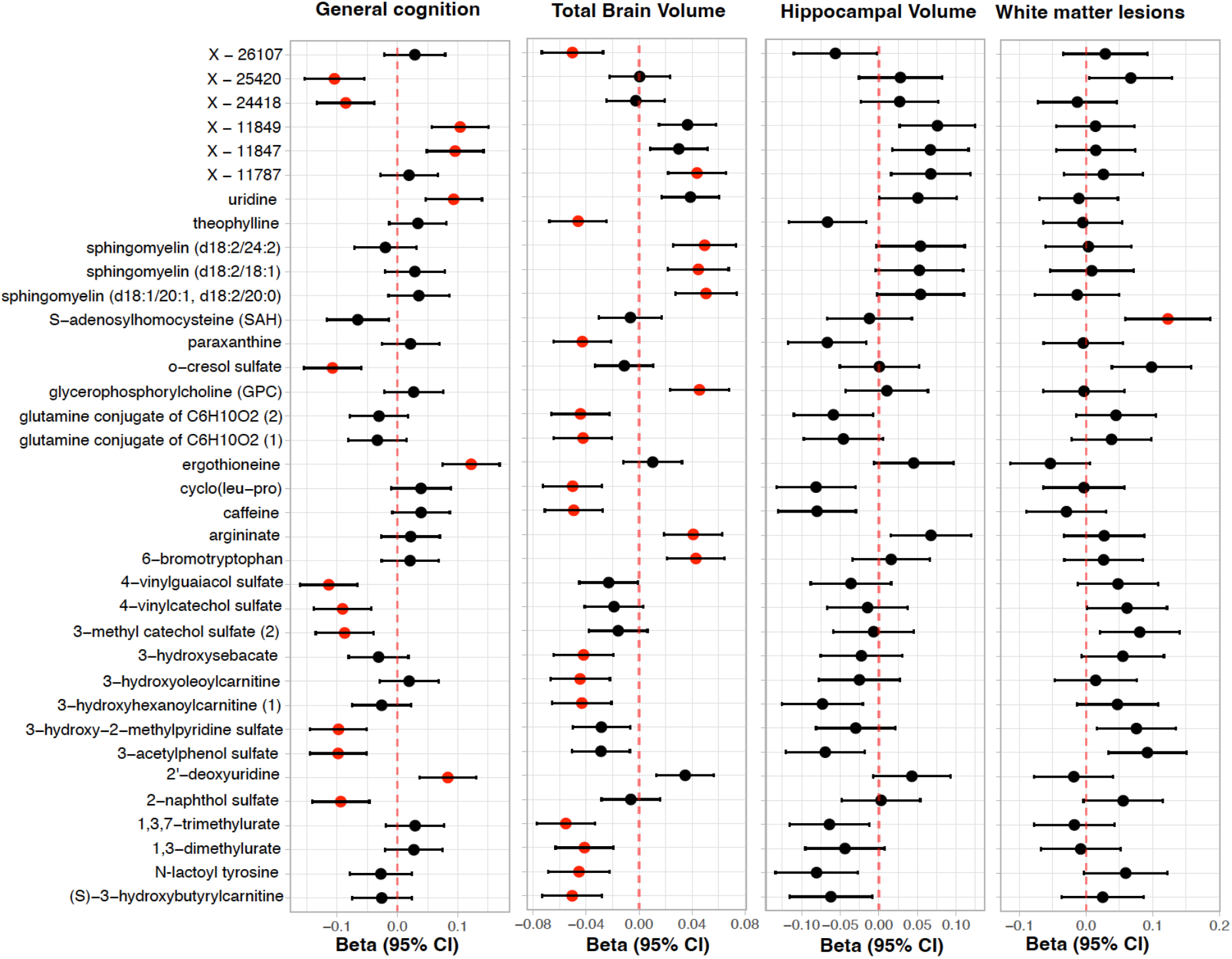
Association of metabolites (from left to right) with general cognition, total brain volume, total hippocampal volume, and total white matter lesions. Red circles indicate beta estimates for metabolites that are significant at FDR < 0.05. Note: N-lactoyltyrosine is the updated annotation of a metabolite that was annotated as 1-carboxyethyltyrosine in the original data set. This correction was provided by Metabolon.

**Table 2:**
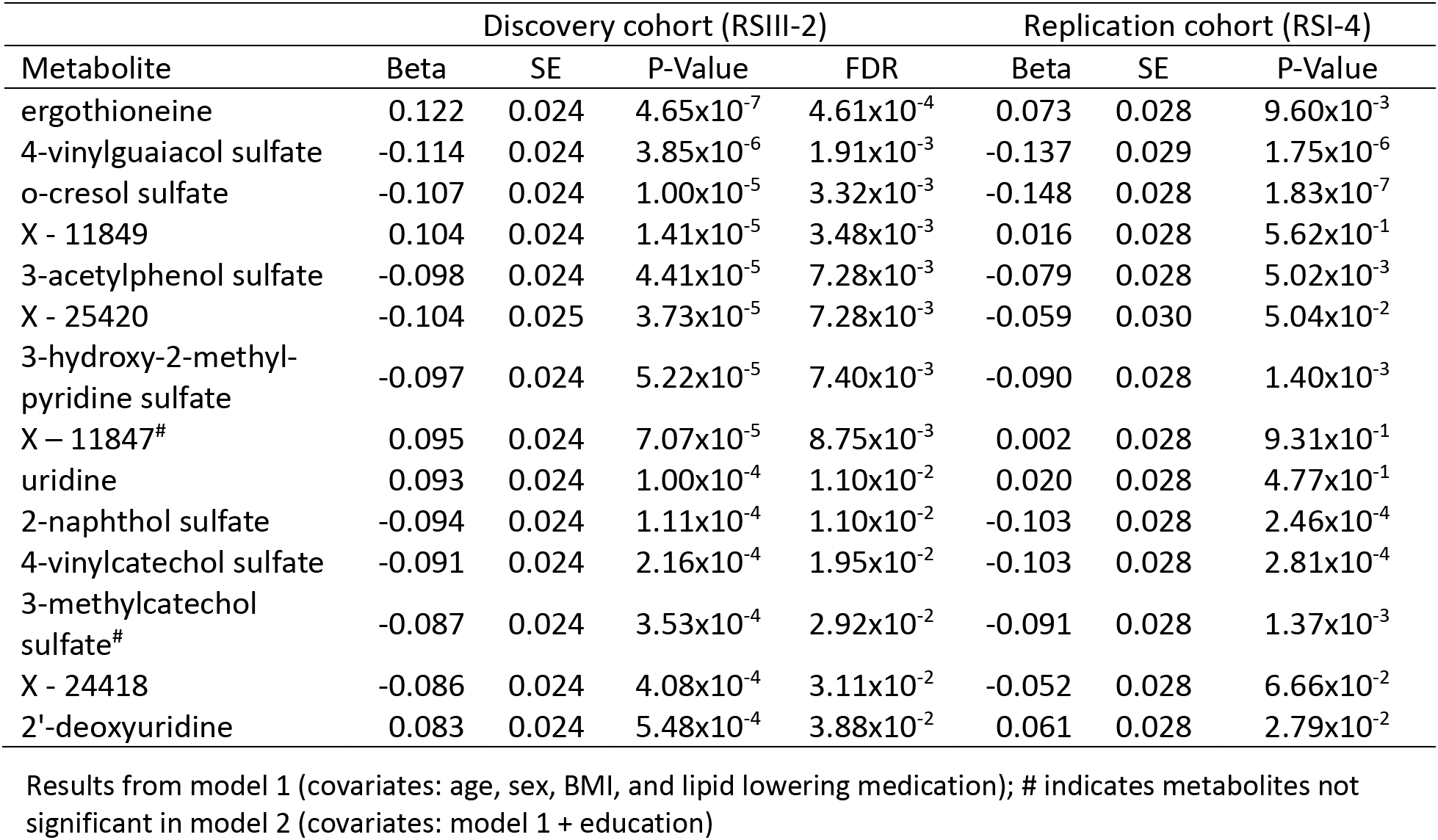
Metabolites associated with general cognition.

| Metabolite | Discovery cohort (RSIII-2) |  |  |  | Replication cohort (RSI-4) |  |  |
| --- | --- | --- | --- | --- | --- | --- | --- |
|  | Beta | SE | P-Value | FDR | Beta | SE | P-Value |
| ergothioneine | 0.122 | 0.024 | $4.65 \times 10^{-7}$ | $4.61 \times 10^{-4}$ | 0.073 | 0.028 | $9.60 \times 10^{-3}$ |
| 4-vinylguaiacol sulfate | -0.114 | 0.024 | $3.85 \times 10^{-6}$ | $1.91 \times 10^{-3}$ | -0.137 | 0.029 | $1.75 \times 10^{-6}$ |
| o-cresol sulfate | -0.107 | 0.024 | $1.00 \times 10^{-5}$ | $3.32 \times 10^{-3}$ | -0.148 | 0.028 | $1.83 \times 10^{-7}$ |
| X - 11849 | 0.104 | 0.024 | $1.41 \times 10^{-5}$ | $3.48 \times 10^{-3}$ | 0.016 | 0.028 | $5.62 \times 10^{-1}$ |
| 3-acetylphenol sulfate | -0.098 | 0.024 | $4.41 \times 10^{-5}$ | $7.28 \times 10^{-3}$ | -0.079 | 0.028 | $5.02 \times 10^{-3}$ |
| X - 25420 | -0.104 | 0.025 | $3.73 \times 10^{-5}$ | $7.28 \times 10^{-3}$ | -0.059 | 0.030 | $5.04 \times 10^{-2}$ |
| 3-hydroxy-2-methyl-pyridine sulfate | -0.097 | 0.024 | $5.22 \times 10^{-5}$ | $7.40 \times 10^{-3}$ | -0.090 | 0.028 | $1.40 \times 10^{-3}$ |
| X – 11847<sup>#</sup> | 0.095 | 0.024 | $7.07 \times 10^{-5}$ | $8.75 \times 10^{-3}$ | 0.002 | 0.028 | $9.31 \times 10^{-1}$ |
| uridine | 0.093 | 0.024 | $1.00 \times 10^{-4}$ | $1.10 \times 10^{-2}$ | 0.020 | 0.028 | $4.77 \times 10^{-1}$ |
| 2-naphthol sulfate | -0.094 | 0.024 | $1.11 \times 10^{-4}$ | $1.10 \times 10^{-2}$ | -0.103 | 0.028 | $2.46 \times 10^{-4}$ |
| 4-vinylcatechol sulfate | -0.091 | 0.024 | $2.16 \times 10^{-4}$ | $1.95 \times 10^{-2}$ | -0.103 | 0.028 | $2.81 \times 10^{-4}$ |
| 3-methylcatechol sulfate<sup>#</sup> | -0.087 | 0.024 | $3.53 \times 10^{-4}$ | $2.92 \times 10^{-2}$ | -0.091 | 0.028 | $1.37 \times 10^{-3}$ |
| X - 24418 | -0.086 | 0.024 | $4.08 \times 10^{-4}$ | $3.11 \times 10^{-2}$ | -0.052 | 0.028 | $6.66 \times 10^{-2}$ |
| 2'-deoxyuridine | 0.083 | 0.024 | $5.48 \times 10^{-4}$ | $3.88 \times 10^{-2}$ | 0.061 | 0.028 | $2.79 \times 10^{-2}$ |

Results from model 1 (covariates: age, sex, BMI, and lipid lowering medication); # indicates metabolites not significant in model 2 (covariates: model 1 + education)

Testing MRI markers for total brain volume, total hippocampal volume, and white matter lesions, we observed significant association between 21 metabolites with total brain volume and one metabolite (S-adenosylhomocysteine (adjusted mean difference = 0.123, *P* = 1.73x10^- 4^) with total white matter lesions. None of the metabolites was significantly associated with total hippocampal volume (**Figure 1, Supplementary Table 2)**. Among the significant metabolites, higher levels of three sphingomyelins, glycerophosphorylcholine (GPC), 6-bromotryptophan, argininate, and X – 11787 were associated with higher brain volume. In contrast, lower levels of caffeine, theophylline, paraxanthine, 1,3,7-trimethylurate, 1,3-dimethylurate, cyclo(leu-pro), (S)-3-hydroxybutyrylcarnitine, 3-hydroxyhexanoylcarnitine, 3-hydroxyoleoylcarnitine, 3-hydroxysebacate, N-lactoyltyrosine, two metabolite signals of glutamine conjugates of C6H10O2 (2), and X – 26107 showed association with higher brain volume. These metabolites can be assigned to one of four main sets of highly correlated metabolites (**Supplementary Figure 3**), with (i) six metabolites related to the caffeine pathway (0.46 < r < 0.92), (ii) three sphingomyelins and glycerophosphorylcholine (GPC) (0.24 < r < 0.77), (iii) seven metabolites including three hydroxylated acylcarnitines and a hydroxylated dicarboxylic fatty acid (0.41 < r < 0.85), and (iv) arginate, N-lactoyltyrosine, X – 11787, and S-adenosylhomocysteine (SAH) (associated with white matter lesions) (0.24 < r < 0.42). 6-bromotryptophan moderately correlated (r = 0.32) with 2-deoxyuridine, which was associated with cognition.

In sex stratified analyses, only one metabolite (4-vinylguaiacol sulfate) was significantly (FDR < 0.05) associated with cognition in males (adjusted mean difference = -0.143, *P* = 2.73x10^-5^), and only three metabolites (1,3,7-trimethylurate, caffeine, sphingomyelin (d18:1/20:1, d18:2/20:0)) showed significant association (FDR < 0.05) with total brain volume in female participants. These findings are most likely explained by the reduced statistical power in the stratified analyses.

### Metabolite signatures of cognition and MRI markers are concordant

While the set of metabolites significantly associated with cognition (n = 14) and MRI markers (n = 22) did not overlap, the patterns of effects as visualized in the forest plots in **Figure 1** suggest similarities in the observed effects. We tested the overall concordance of metabolite signatures across these related phenotypes by assessing the correlation of the regression coefficients for cognition and MRI markers, including all metabolites that showed nominal association (*P* < 0.05) to one of the compared phenotypes. Regression coefficients for the metabolite associations of cognition significantly correlated with the coefficients from the association with total brain volume (**Supplementary Figure 4**B; r = 0.33, *P* = 4.66x10^-11^), with total hippocampal volume (**Supplementary Figure 4**C; r = 0.27, *P* = 1.15x10-7), and with white matter lesions (**Supplementary Figure 4**D; r= -0.49, *P* = 5.39x10^-24^).

Out of 105 metabolites that nominally associated (*P* < 0.05) with cognition, 49 also showed nominal association to at least one of the three MRI phenotypes (**Supplementary Figure 4**A). 3-acetylphenol sulfate was the only metabolite that showed significant association with general cognition (adjusted mean difference = -0.098, *P* = 4.41x10^-5^) and nominal association with all three MRI phenotypes (total brain volume (-0.029, *P* = 9.59x10^-3^), hippocampal volume (-0.069, *P* = 7.89x10^-3^), white matter lesions (0.092, *P* = 2.24x10^-3^)). Moreover, out of the 14 metabolites significantly associated with cognition, concordant effects were observed for ergothioneine between cognition, hippocampal volume, and white matter lesions, for uridine, 2’-deoxyurdine, X – 11847, and X – 11849 between cognition, hippocampal volume, and total brain volume but not white matter lesions. Conversely, the uncharacterized metabolites X – 24418 and X – 25420 did not show any effects in the association with MRI phenotypes.

### Metabolite associations with cognition replicate in an independent cohort and overlap with AD metabolite signature

We replicated the findings obtained from the association of metabolites with general cognition in this study (RSIII-2 cohort) in participants from the RS1-4 cohort. Among the 14 metabolites associated with general cognition (FDR < 0.05), nine metabolites showed significant association (*P* < 0.05) in the replication analysis with concordant direction of association (**Table 2**).

We additionally compared the regression coefficients of metabolites associated with general cognition in RSIII-2 (*P* < 0.05) to the regression coefficients observed for the association of these metabolites with incident AD diagnosis in the RSI-4 cohort and found a significant correlation between the signals (r = -0.73, *P* = 1.03x10^-13^) (**Figure 2**, **Supplementary Table 3**). Among the cognition-related metabolites (*P* < 0.05), beta-cryptoxanthin and imidazole propionate showed the largest effects with incident AD diagnosis (beta-cryptoxanthin: -0.210, *P* = 2.54x10^-4^; imidazole propionate: 0.199, *P* = 5.11x10^-4^).

**Figure 2:**
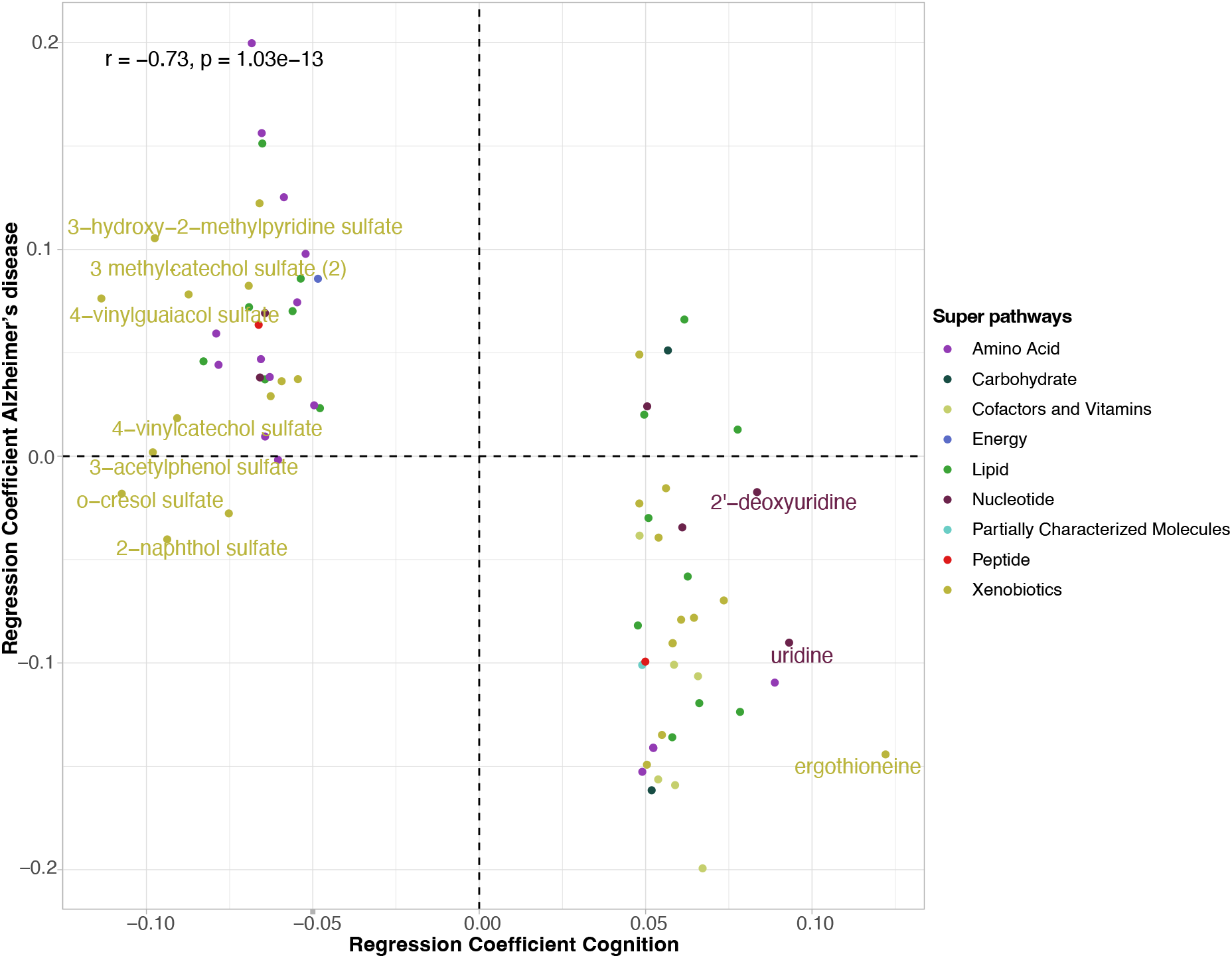
Concordance of metabolite signatures between cognition in the RSIII-2 cohort and incident AD diagnosis in the older RSI-4 cohort, comparing the regression coefficients of metabolites nominally associated (*P* < 0.05) with general cognition with their coefficients for incident AD (cox regression). Colors indicate the classes of metabolites.

### Exposome has strongest influences on metabolites associated with cognition and MRI markers

To investigate which unmodifiable and potentially modifiable factors influence the blood levels of the 36 metabolites that associated with general cognition or MRI markers, we estimated how much of each metabolite’s variance is explained by genetic (single nucleotide polymorphisms (SNPs)), lifestyle (BMI, alcohol consumption, smoking, and education), clinical (diabetes, blood pressure), medication (n = 31), and microbial features (based on 16S rRNA sequencing data) in the RSIII-2 cohort (see **Methods; Supplementary Table 4**). For genetic factors, we chose a conservative approach by only considering SNPs that showed genome-wide significant associations (*P* = 5.0x10^-8^) with metabolite levels in the RSIII-2 cohort. For comparison with genetics-based explained variance (EV), SNP-based heritability estimates are provided in **Supplementary Table 5**.

Among the metabolites associated with general cognition, lifestyle features explained considerable parts of the variance of nine metabolites, including 2-naphthol sulfate (28.6 %), o-cresol sulfate (21.0 %), 4-vinylguaiacol sulfate (11.3 %), 4-vinylcatechol sulfate (7.8 %), and ergothioneine (4.7 %) (**Figure 3A**). Genetics explained 1.2 % of the variance of 2-deoxyuridine and 0.9 % of X – 24418. While clinical factors did not account for the variance observed for any of the 14 cognition-associated metabolites, medication did explain part of the variance for six metabolites, with the highest value being observed for ergothioneine (3.6 %). Gut-microbiota also explained part of the variance for six of the cognition-associated metabolites, including 3-hydroxy-2-methylpyridine sulfate (5.4 %), 4-vinylguaiacol sulfate (4.3 %), and ergothioneine (3.9 %). Interestingly, we observed higher influence of the gut microbiome for the metabolites that showed largest effect sizes in the analysis of incident AD diagnosis in the older RSI-4 cohort. For example, the gut microbiome explained more than 5 % of the variance in measured blood levels (**Supplementary Tables 3and 4**, **Supplementary Figure 5**) of various bile acids and the cognition-related metabolites (*P* < 0.05) imidazole propionate (11.3 %) and beta-cryptoxanthin (5.1 %).

**Figure 3:**
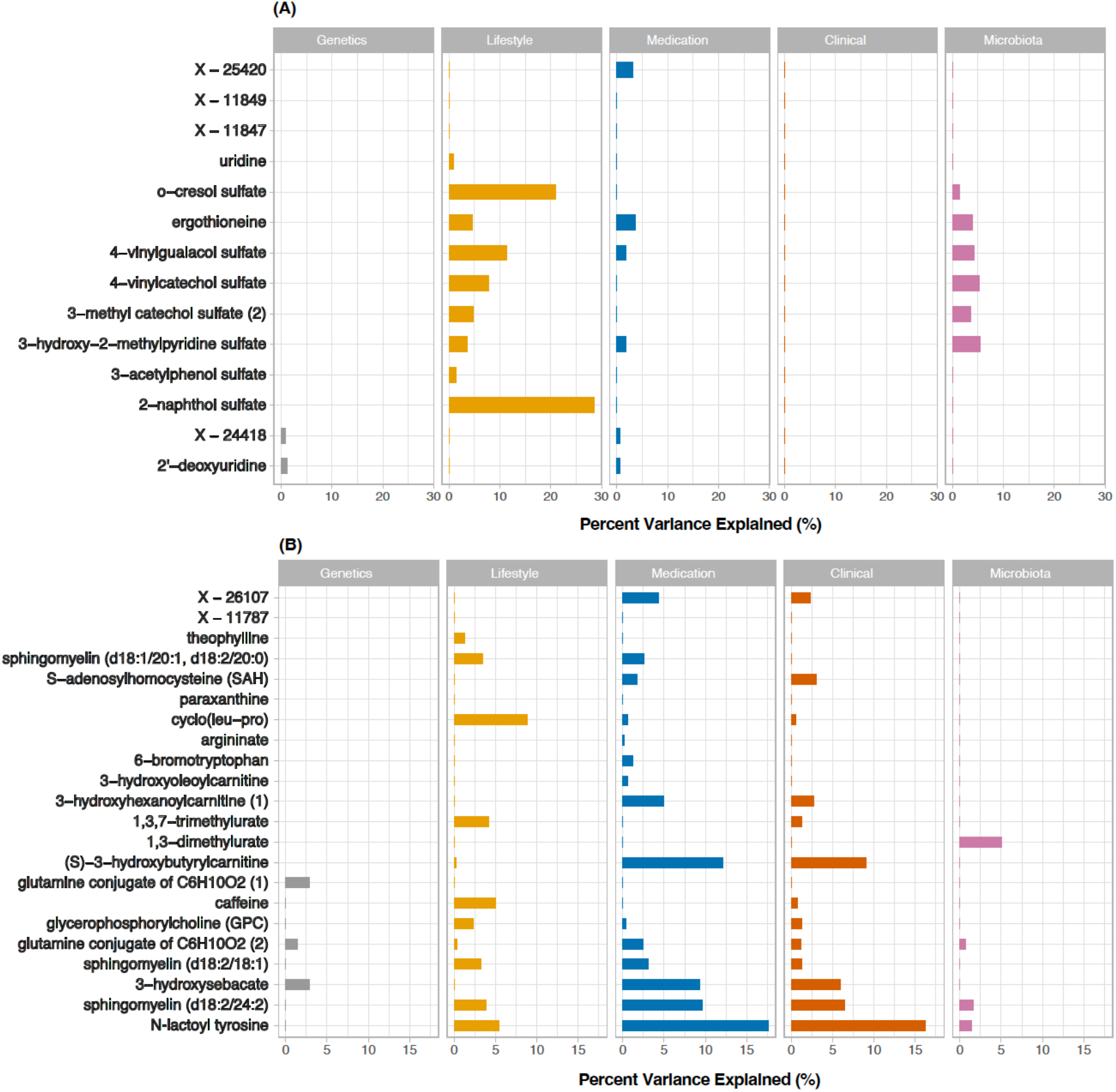
Explained variance (EV) of general cognition-(A) and MRI marker-associated (B) metabolites by genetics, microbiota, lifestyle, clinical variables, and medication use. Note: N-lactoyltyrosine is the updated annotation of a metabolite that was annotated as 1-carboxyethyltyrosine in the original data set. This correction was provided by Metabolon.

For metabolites associated with total brain volume or white matter lesions, lifestyle (11 metabolites, EV range 0.20 – 8.8 %), medication use (15 metabolites, range 0.20 – 17.7 %) and clinical factors (13 metabolites, EV range 0.6 – 16.2 %) were the major factors influencing the metabolites’ variance in our study. Gut microbiota explained part of the variance of four metabolites (EV range 0.7 – 5.1 %). Genetic factors explained some of the variance of three metabolites (EV range 1.5 – 2.9 %) (**Supplementary Table 4**, **Figure 3B**).

Next, we compared the influences on the cognition- and MRI-related metabolites with those observed for all profiled metabolites (**Figure 4**) and tested whether the associated metabolites were enriched in the set of metabolites with EV > 0 for any factor. Out of the 991 tested metabolites, genetics explained some variance (maximal 67.0 %) for 130 metabolites. Gut-microbiota contributed to the levels of 148 metabolites, explaining up to 34.7 % of the variance. Lifestyle factors explained up to 29.6 % of the variance for 333 metabolites, clinical features explained up to 18.4 % for 188 metabolites, and medication use explained up to 40.3% for 430 metabolites. Among the five tested feature groups, EV values (EV > 0) of metabolites for clinical features showed a strong positive correlation (FDR < 0.05) with those for medication use (r = 0.660) and lifestyle factors (r = 0.202) (**Supplementary Figure 6**).

**Figure 4:**
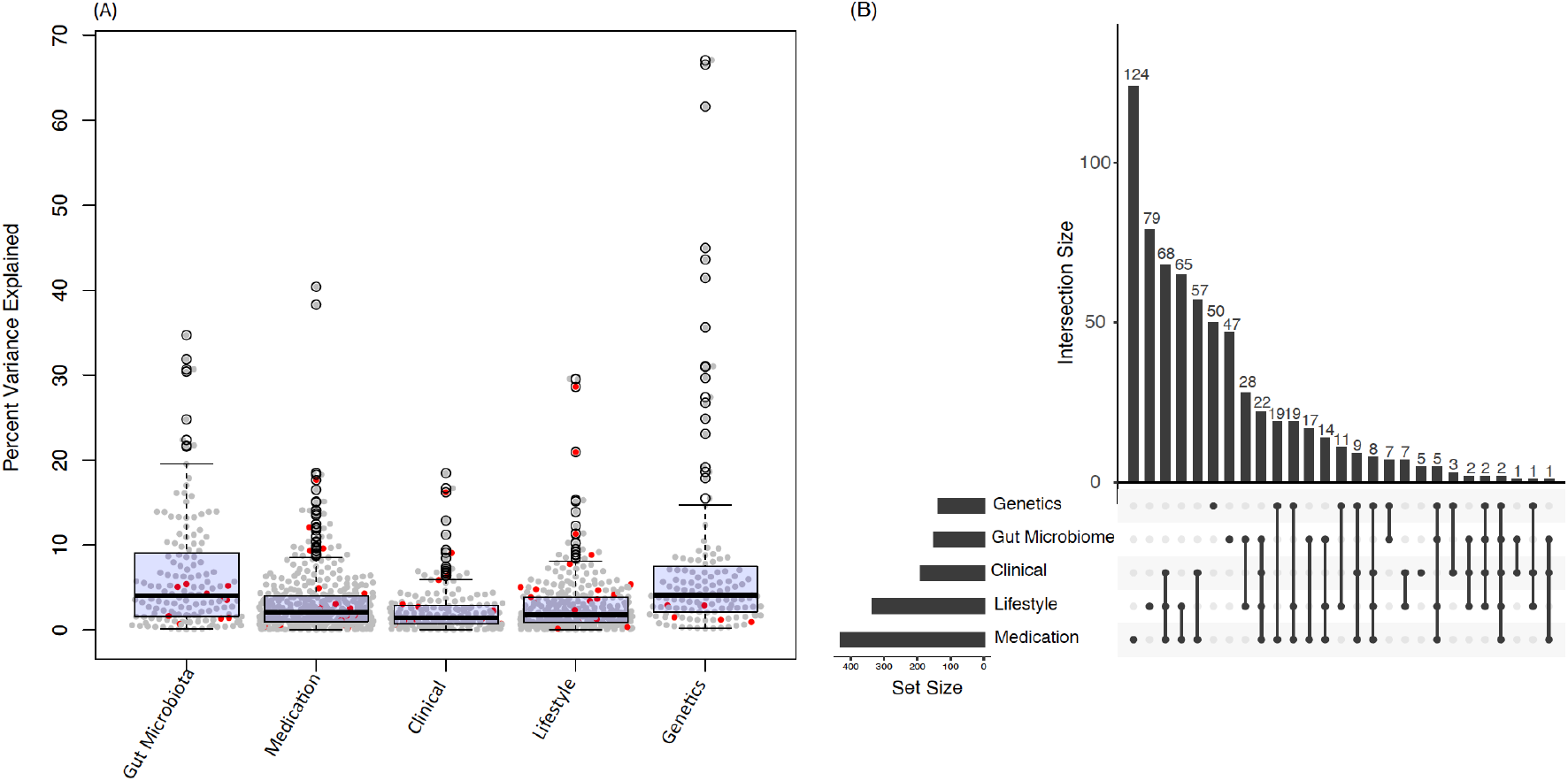
Explained variance of metabolites by gut microbiome, medication use, clinical factors, lifestyle, and genetics (A). Red dots represent metabolites associated with general cognition or with magnetic resonance imaging markers (FDR < 0.05). (B) Overlap of metabolites explained (EV > 0) by the five tested classes of features.

For the 36 cognition- and MRI-related metabolites, significant enrichment (considering the EVs of all 991 studied metabolites) was observed for the influence of lifestyle factors and clinical features, with 20 metabolites being explained to some extent by lifestyle factors (*P* = 6.48x10^-3^) and 13 metabolites by clinical features (*P* = 1.48x10^-2^). We did not observe significant enrichment for medication use or genetics (medication use: 21 metabolites, *P* = 8.56x10^-2^; genetics: 5 metabolites, *P* = 0.803). With 10 cognition- or MRI-related metabolites, for which the variance was partially explained by gut microbiota, the enrichment narrowly missed the significance threshold (*P* = 5.15x10^-2^).

### Associations of cognition- and MRI-associated metabolites with individual level features

To further disentangle the determinants of metabolites associated with cognition and MRI phenotypes, we checked their association with lifestyle factors, clinical features, medication use, and gut microbiota in univariate regression analyses. The complete results are provided in **Supplementary Tables 6 - 9.**

Out of the 14 metabolites associated with general cognition, 13 metabolites showed significant association (FDR < 0.05) with different lifestyle factors (**Supplementary Figure 7, Supplementary Table 6)**. In particular, smoking was associated with higher levels of the sulfated metabolites (2-naphthol sulfate, o-cresol sulfate, 3-hyroxy-2-methylpyridine sulfate, 3-methylcatechol sulfate, 4-vinylcatechol sulfate, 4-vinylguaiacol sulfate, 3-acetylphenol sulfate) and X – 25420 and with lower levels of uridine and 2’-deoxyuridine (i.e., matching the association pattern of worse cognition). Ergothioneine was associated with all tested factors except smoking; thereby, higher blood levels of ergothioneine, which were linked to better cognition, were associated with higher alcohol intake and higher education, and with lower BMI.

Among the tested clinical factors and medications, diabetes and antidiabetic medication were the factors with the highest number of associations (**Supplementary Figure 7**, **Supplementary Tables 7 and 8**). Higher levels of five out of the seven sulfates (including 3-methylcatechol sulfate) and lower levels of 2’-deoxyuridine were significantly associated with diabetes and, with the exception of o-cresol sulfate, also with antidiabetic medication. Interestingly, higher levels of 3-methylcatechol sulfate, which were associated with worse cognition, were associated with lower systolic and diastolic blood pressure. Conversely, higher levels of ergothioneine linked to better cognition were associated with lower blood pressure. Antacids, thyroid therapy, and psychoanaleptics were associated with lower blood levels of ergothioneine, with largest effect being observed for antacid use (-0.45, *P* = 4.27x10^-9^).

While smoking was the major lifestyle factor associated with metabolites linked to cognition, alcohol intake and BMI showed the highest number of associations observed for the metabolites linked to MRI phenotypes, with 20 out of the 22 metabolites being associated to BMI and/or alcohol intake (**Supplementary Figure 8, Supplementary Table 6)**. Thereby, higher levels of seven metabolites, of which six are known to be linked to coffee intake and the caffeine pathway (e.g., cyclo(leu-pro), caffeine, theophylline, 1,3-dimethylurate), and lower levels of five metabolites (sphingomyelin (d18:2/18:1), S-adenosylhomocysteine (SAH), argininate, N-lactoyltyrosine, X – 11787) were associated with higher alcohol intake. Six metabolites that were associated with alcohol intake were also associated with BMI. Thereof three metabolites were associated in the opposite effect direction (N-lactoyltyrosine, argininate, SAH) and three coffee-related metabolites in the same direction. Regarding clinical factors and medication, most associations were observed for diabetes, hypertension, and antidiabetic medication (**Supplementary Figure 8, Supplementary Tables 7 and 8)**. The association patterns largely overlapped and resembled the effect directions of worse brain health and higher BMI, i.e., the patterns were characterized by lower levels of sphingomyelins (sphingomyelin (d18:2/24:2)) and GPC and higher levels of N-lactoyltyrosine, SAH, X – 26107, and acylcarnitines carrying hydroxylated lipids (e.g., 3-hydroxyhexanoylcarnitine). Except five metabolites linked to the caffeine pathway, all MRI-associated metabolites were associated with some medication, of which the association of antidiabetic therapy and N-lactoyltyrosine showed the lowest *P*-value (adjusted mean difference = 1.54, *P* = 2.84x10^-36^). Three of these metabolites were influenced by seven or more drug types ((S)-3-hydroxybutyrylcarnitine, sphingomyelin (d18:2/24:2), N-lactoyltyrosine), with N-lactoyltyrosine showing associations with antidiabetic, thyroid, and cardiac therapies, statins and other lipid lowering agents, ACE inhibitors, betablockers, diuretics, calcium blockers, and antithrombotic agents.

Out of the 36 metabolites associated with general cognition and MRI phenotypes, 22 metabolites showed significant association (FDR < 0.05, considering all profiled metabolites (n = 991) and microbial features) with specific gut microbiota (**Supplementary Table 9**, **Supplementary Figure 9**). Ergothioneine showed significant associations with 12 microbial genera. Increased levels of ergothioneine were associated with a higher abundance of *genus Lachnospiraceae ND3007* ()*, genus NK4A214, genus Fusicatenibacter, genus Romboutsia, genus Erysipelotrichaceae UCG-003* and *genus UCG-003*. Moreover, higher abundance of *genus DTUO89* was associated with higher levels of sulfate-containing metabolites in our study (3-acetylphenol sulfate, o-cresol sulfate, 4-vinylguaiacol sulfate, 4-vinylcatechol sulfate, 3-methylcatechol sulfate, 3-hydroxy-2-methylpyridine sulfate). Also, higher abundance of *genus Lachnospiracea UCG-010* showed association with lower levels of three of the sulfated metabolites (3-acetylphenol sulfate, 4-vinylcatechol sulfate, and 3-hydroxy-2-methylpyridine sulfate). Among the MRI marker-associated metabolites, higher levels of (S)-3-hydroxybutyrylcarnitine, 3-hydroxyhexanoylcarnitine, and two glutamine conjugates of C6H10O2 were associated with increased abundance of genus *Ruminococcus Torques* group. Likewise, higher levels of sphingomyelins were linked to higher abundance *of genus Clostridium sensus Stricto 1*, *genus Coprococcus*, and *genus Faecalibacterium*, and lower abundance of *genus Veillonella, genus Rothia, genus Atopobium, genus Phocea, genus Sellimonas, genus UC5-1-2E3.* Levels of the caffeine-related metabolites (caffeine, 1,3,7-trimethylurate 1,3-dimethylurate, paraxanthine, theophylline) were associated with 16 microbial genera (**Supplementary Figure 9)**.

### Ergothioneine mediates the association between antacid medication and cognition

Ergothioneine robustly associated with general cognition and incident AD and was influenced by features of various factors of the exposome and the gut microbiome (**Supplementary Figure 10**). In particular, we found a strong effect for the association between antacid medication and the blood levels of this metabolite. Considering recent reports of associations between the use of proton pump inhibitors (PPIs), a common class of antacid medication, and dementia, we investigated the potential mediating role of ergothioneine. As result of the mediation analysis performed in the RSIII-2 cohort, we found evidence for 31.5 % (CI: 15.5 – 71 %) of the total effect of antacid medication on cognition (-0.235, *P* < 2x10^-16^) being mediated by ergothioneine (**Figure 5**, **Supplementary Table 10**).

**Figure 5:**
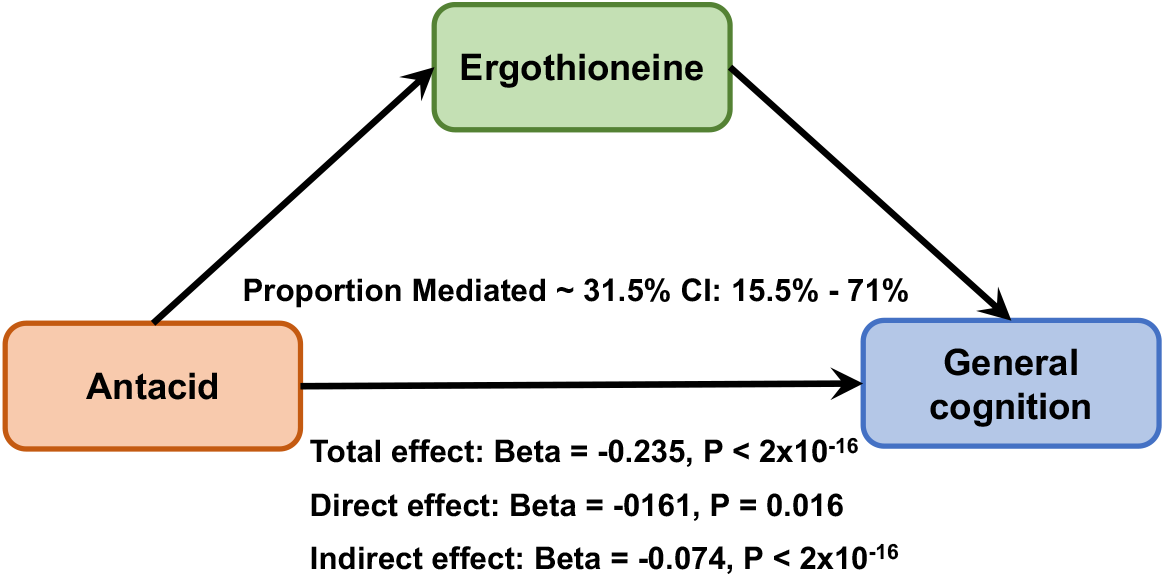
Mediation analysis for the relationship of antacid use, ergothioneine, and general cognition.

As a second example, we further investigated our findings regarding the seven sulfated xenobiotics that associated with general cognition and smoking status. As expected, blood levels of those xenobiotics were consistently higher among current smokers (n = 180) compared to never (n = 350) and former smokers (n = 574) (**Supplementary Figure 11**). Stratification of the association between general cognition and these metabolites based on smoking status showed that the associations for 3-methylcatechol sulfate, 3-vinylcatechol sulfate, 3-acetylphenol sulfate, and 3-hydroxy-2-methylpyridine sulfate remained significant in the group of current smokers. 4-vinylcatechol sulfate, 4-vinylguaiacol sulfate, and o-cresol sulfate showed significant associations with cognition in former smokers. In case of 3-hydroxy-2-methylpyridine sulfate, of which the variance was more explained by gut microbiota (EV 5.4%) than by lifestyle (3.7 %), the association with cognition was still significant within the stratum of never smokers.

## DISCUSSION

In our study, we observed significant association of 14 metabolites with general cognition and 22 metabolites with MRI markers in participants without dementia. Nine of the 14 metabolite associations with cognition were replicated in an independent dementia-free sample, which included older participants. Thereby, the effects that we saw for cognition correlated significantly with those of incident AD diagnosis in the replication cohort (RSI-4) and with effects for the imaging markers (total brain volume and hippocampal volume) in the discovery cohort (RSIII-2), confirming that the metabolite signature derived for cognition in participants prior to the onset of any symptoms of dementia is of relevance to understand early metabolic changes in AD etiology.

In the investigated population-based cohort, the variance of the circulating metabolites associated with cognition or MRI markers were mainly driven by lifestyle and clinical factors as well as medication use. Genetics and the gut microbiome explained less of the variance for these metabolites in our discovery cohort. To further disentangle the complex relationship of the cognition- and MRI marker-associated metabolites with potentially modifiable factors of the exposome or the gut microbiome, we provide a thorough overview of metabolite associations with the single lifestyle, medication, clinical, and gut microbial features available in our cohort. Integrating the information of association studies on AD-relevant endophenotypes with associations related to metabolite origin may improve our understanding of disease etiology and may help to prioritize and identify targets for prevention, as demonstrated for selected examples in the following.

Ergothioneine, a naturally occurring, sulfur-containing amino acid, showed the strongest association with cognition with higher blood levels relating to better general cognition in our study. Considering the previously reported cytoprotective function of ergothioneine (30), this result suggests a neuroprotective role for this metabolite. While little is known about the transport of ergothioneine from blood to brain, recent studies based on brain samples showed that lower levels of ergothioneine in brain associated with worse cognition, cognitive decline, and AD diagnosis (31, 32). In our study, we observed that gut microbiota, medication use, and lifestyle factors explain part of the inter-individual variation of ergothioneine in blood. As dietary data was not available for the RSIII-2 samples, we were not able to assess the influence of food directly. However, ergothioneine has been previously reported to be mainly derived from external food sources (including mushrooms) (33), with diet explaining around 14% of the variation of ergothioneine (25). In addition to diet, several factors may contribute towards the levels of ergothioneine in blood, including age (34), and genetics (35). In our study, we newly found significant associations of antacids, psychoanaleptics, thyroid therapy, systolic and diastolic blood pressure with lower levels of ergothioneine. In contrast, alcohol intake and higher education were found associated with higher levels of this metabolite in blood. When investigating the relationship of antacid intake with ergothioneine levels further, we found evidence for a mediating role of ergothioneine in the association of antacid with general cognition in our data. Antacids such as proton pump inhibitors are commonly prescribed medications to lower acid concentration in the stomach (36). For proton pump inhibitors, various observational studies reported negative associations with phenotypes of neurodegenerative diseases (37-39). Our results suggest that the reduction of ergothioneine as one possible mechanism explaining harmful effects of this medication on neurodegenerative diseases. Thereby, it remains unclear whether these medications affect the absorption of ergothioneine from diet or whether they influence the gut microbiome which may have influence on ergothioneine levels in blood. In our study, we also observed significant associations of six microbial genera with lower levels of ergothioneine and 6 microbial genera with higher blood levels of ergothioneine. Among the genera whose higher abundance were associated with lower levels of ergothioneine include genus *Clostridium innocuum group, Sellimonas, Hungatella, Eisenbergiella, Enterobacter, Flavonifractor.* Among these six genera, five belong to the phylum Firmicutes whose increased abundance has been reported in mild cognitive impairment (MCI), the prodromal phase of AD (40)*. Clostridium innocuum* is a pathogenic bacteria associated with western high-fat and high-sugar diet (41). Higher abundance of *genus Sellimonas* and *Hungatella* have been linked to depression (42), and *Sellimonas* to atherosclerotic cardiovascular disease (43), whereas both depression and cardiovascular disease are risk factors for AD and cognitive decline (44, 45). Further evidence suggests that infection with *Enterobacter* may contribute to progression of neurodegeneration (46, 47). Moreover, we found an inverse association of *Enterobacter* with glycerophosphorylcholine, of which higher levels were associated with higher brain volume in our study. In summary, these pieces of evidence point towards an important role of ergothioneine for maintaining cognitive function and highlight the potential of intervention and prevention schemes targeting the management of ergothioneine.

As a second example, lower levels of seven sulfated xenobiotics, including o-cresol sulfate, were associated with better general cognition in our study. Out of these metabolites, three sulfates also showed association with MRI phenotypes. In general, o-cresols, which are related to the metabolism of toluene as contained in smoke, glues, and cleaners, are very specific biomarkers for toluene exposure (48, 49). Thus, not surprisingly, the variance in the blood levels of o-cresol sulfate (and most of the correlated sulfates except 3-acetylphenol sulfate and 3-hydroxy-2-methylpyridine sulfate) was mostly explained by lifestyle factors, with smoking being the main driver of these metabolites and their association to cognition in our study, as shown in a sensitivity analysis. However, interestingly, the two sulfated, smoking-associated metabolites 4-vinylguaiacol sulfate and 4-vinylcatechol sulfate, which weakly correlated with the MRI-associated coffee-related metabolites, showed association with cognition both in former and current smokers. Also, the association of 3-hydroxy-2-methylpyridine sulfate with general cognition remained significant in never smokers. These findings suggest relevance of those metabolites for AD pathogenesis beyond reflecting smoking behavior, which has been recognized as an important modifiable risk factor of AD previously (50, 51).

N-lactoyltyrosine, which belongs to a class of pseudopeptides, formed by lactic acid and an amino acid (52), was associated with brain health in our study, with higher levels being linked to lower brain volume. N-lactoyl amino acids received some attention in diabetes research recently (53, 54); while higher levels of N-lactoyl amino acids (including N-lactoylphenylalanine, N-lactoyltyrosine, and N-lactoylleucine) associate with decreasing glycemic control and incident diabetes, for N-lactoylphenylalanine it has been shown that it reduces appetite in mice (53) and mediates the effect of metformin on appetite and weight reduction in humans (55, 56). Also, in the RSIII-2 cohort, the N-lactoyl amino acids are the metabolites most strongly associated with antidiabetic therapy, confirming these previous findings. Besides N-lactoyltyrosine, further metabolites that associated with MRI markers, such as glycerophosphorylcholine and hydroxylated acylcarnitines, resembled the association patterns observed for diabetes and BMI despite adjustment for BMi in the model, emphasizing the shared risk profiles between diabetes and AD. Interestingly, the typical markers of glycemic control such as glucose or 1,5-anhydroglucitol were not found associated with MRI markers in our study.

While the gut microbiome was not prominent in explaining the variance of the cognition and MRI marker-associated metabolites, some of the metabolites that were mainly driven by gut microbiota have been found associated with AD or AD risk previously (57-62), including bile acids (e.g., deoxycholate) and indole-derived metabolites (e.g., indoleacetate), or with incident AD in the RSI-4 replication cohort, in particular, imidazole propionate and beta-cryptoxanthin. In the younger RSIII-2 discovery cohort, these metabolites showed nominal significance with general cognition but did not survive correction for multiple testing. These observations suggest that gut microbiome-related metabolites might play a more important role later in the disease process. Nonetheless, the generally observed good concordance of metabolite effects related to cognition in the younger RSIII-2 cohort with the metabolite signature of incident AD diagnosis in the older RSI-4 cohort in our study, highlights the involvement of the gut microbiome and the identified microbiome-related metabolites also in asymptomatic phases of AD development, suggesting these metabolites and their related microbiota as promising targets for modification through interventions.

### Limitations

While building on rich data from a population-based cohort, our study has several limitations. First, dietary information was not available for the time of blood collection. As diet is known to be an important lifestyle factor defining the levels of many metabolites, this lack of data clearly limits the insights about the impact of the exposome. Also, the impact of physical activity could not be investigated in this study. While the MRI measurement was also not available exactly at the time of blood and feces collection, we addressed this issue by adjusting for the time difference in our analyses. Second, the estimation of explained variance strongly depends on the features used as input. For example, applying a less strict threshold for the inclusion of genetic variants will result in overall higher estimates for the genetic contribution for all metabolites. With the chosen approach the EVs for genetics were similar to those previously reported by Bar *et al.* (25). Also, metabolites driven by gut-microbiota showed similar number of metabolites between our study (n = 148) and Bar *et al*. 2020 (n = 182) and similar EV percentages (e.g., cinnamoylglycine (current study = 24% versus Bar et al., 24%), 3-phenylpropionate (31 % versus 26 %), p-cresol sulfate (32 % versus 42 %), and p-cresol glucuronide (22 % versus 41 %) (25). Differences in the EV of metabolites by lifestyle factors and clinical variable with earlier studies (25, 63) can be attributed to the difference in the inclusion of various features in these types of predictors. Finally, clinical features and medication use are inherently connected. As a consequence, we cannot differentiate between the effects of the two factors on metabolite levels within the setting of a population-based cohort as used in our work. In general, we cannot infer causality through any of the analyses performed in this study.

## Conclusion

In conclusion, our study provides compelling evidence that, lifestyle factors play a major role in shaping blood metabolites associated with general cognition and MRI markers in dementia-free participants. The overall concordance of the associations between metabolites and cognition with metabolite signatures of incident AD in an independent sample pinpoints the relevance of the identified metabolites for the disease. Among the potential determinants of these metabolites, smoking emerged as an important lifestyle factor affecting plasma levels of metabolites associated with cognition, while alcohol intake, BMI, diabetes and diabetes medication were linked to MRI marker-associated metabolites. With the example of antacid medication, for which we showed a negative effect on the blood levels of the presumably neuroprotective metabolite ergothioneine, we demonstrated the potential of our approach and the here derived association catalogues for studying the complex interplay between cognition and MRI phenotypes with metabolism and its influences through genetics, gut microbiome, and exposures (lifestyle, medication) to come up with new targets and strategies for disease prevention and early interventions.

## METHODS

### Study population

#### Rotterdam Study

The Rotterdam Study (RS) is a prospective population-based study located in the Ommoord district of Rotterdam, The Netherlands. In 1990, the study was initiated with the inclusion of 7,983 subjects aged 55 years or older (RS-I). From 2000 to 2001, the cohort was expanded with the addition of 3,011 participants ≥ 55 years of age (RS-II). The cohort was further extended with the inclusion of 3,932 participants with age 45 years or older during 2006-2008 (RS-III). All study participants were extensively interviewed and physically examined at their baseline visits and after every 3 to 6 years. The study has been approved by the Medical Ethical Committee of Erasmus Medical Center and by the Ministry of Health, Welfare, and Sport of the Netherlands. Written Informed consents were also obtained from each study participant to participate and to collect information from their treating physicians (64). In the current work, we included data from participants of the second follow-up of the RS-III cohort (RSIII-2) for which gut microbiota, metabolomics, and genetic data were available. We replicated our findings of general cognition in the fourth follow-up of the RS-I cohort (RSI-4).

#### Assessment of lifestyle, clinical factors, and medication intake

In the RS cohorts, information about lifestyle, clinical factors, and medication intake was collected using structural interviews, medical records, and pharmacy data during multiple visits. Information about lifestyle factors such as smoking, alcohol consumption, and educational attainment was collected based on structured home interviews. Smoking data was classified as never, former, or current smokers. Educational attainment was assessed at the baseline visit of the RS cohort and categorized into four groups based on the UNESCO classification: (1) primary education, (2) lower/intermediate general education or lower vocational education, (3) intermediate vocational education or higher education, and (4) higher vocational education or university level (65). In our study, we combined the education categories one and two into primary education. Alcohol consumption was assessed as part of dietary interviews. Alcohol intake in grams per day was calculated based on the number of drinks multiplied by the average amount of ethanol in one drink of the alcoholic beverage. Details of alcohol consumption assessment are described elsewhere (66). Body mass index (BMI) was calculated based on height and weight (kg/m^2^) which were assessed in participants in standing positions without shoes and heavy outer garments. Medical history (clinical factors) and medication intake were compiled based on various sources, including general practitioner records, pharmacy prescription records or a physical examination at the study center. Blood pressure was recorded at the time of the patients’ visit to the study center at the right upper arm in a seated position; the mean of two measurements was recorded. Glucose levels were measured after overnight fasting (8–14 h); diabetes was defined as fasting serum glucose levels ≥ 7.0 mmol/L, non-fasting serum glucose levels ≥ 11.1 mmol/L, and/or the use of antidiabetic medication (ATC-code A10) (67).

#### Genotyping and imputations

Blood from the RS participants was collected during the baseline visit of RS-III. DNA was extracted from blood and genotyping was performed using the 550K, 550K duo, or 610K Illumina arrays. During the genotyping quality control for genetic variants, we applied exclusion criteria, including call rate < 95%, Hardy-Weinberg equilibrium *P* < 1.0x10^-6^, and Minor Allele Frequency (MAF) < 1%. Sample exclusion criteria included excess autosomal heterozygosity, call rate < 97.5%, ethnic outliers, and duplicates or family relationships. Genotypes were imputed using the Markov Chain Haplotyping (MACH) package and the minimac software (68) to the 1000 genome phase 1 version 3 reference panel (69). Among the 1,068 participants with metabolomics data (after preprocessing), genotyping information was available for 925 participants.

#### Metabolomics profiling

We profiled blood plasma samples of 1,082 participants of the RS-III cohort (second follow-up) using the untargeted Metabolon HD4 platform. The resulting data set includes 1,387 metabolites of different classes (lipids, amino acids, xenobiotics, nucleotides, cofactors and vitamins, peptides, carbohydrates, energy-related metabolites, and uncharacterized metabolites). The details of the analytical methods and data extraction procedure of the Metabolon HD4 approach has been described elsewhere and is briefly summarized in **Supplementary Materials**. Based on the batch-normalized data as provided by Metabolon, following additional preprocessing steps were performed: First, 14 participants for which the proportion of missing values across metabolites was greater than 5 times the standard deviation (SD) of the mean missingness in all participants were excluded. Then, metabolites with missingness greater than 70 % were excluded. For the remaining metabolites, the coefficient of variance (CV) of the 64 aliquots of the NIST Standard Reference Material (SRM) 1950 sample, which were measured throughout the experiment, was determined and metabolites with CV greater than 30% were excluded, leaving 1,111 metabolites after the quality control steps. For the present work, we only used data on the 991 frequent metabolites (missingness less or equal 30 %). After log2 transformation, we impute the missing values in our metabolomics data set by applying the K-nearest neighbor (KNN) based imputation method which has been shown to provide robust imputation for metabolomics data previously (70). A detailed flowchart of quality control and preprocessing steps and percentage of major classes of metabolites is provided in the flowchart diagram in **Supplementary Figure 1**.

#### Gut microbiome profiling

Detailed information regarding the collection of fecal samples in the RS-III cohort and the subsequent sequencing procedures have been described previously (71). This sequence data was subjected to a new 16S rRNA profiling pipeline. In short, raw reads were demultiplexed using a custom script to separate sample fastq files based on the dual index. Primers, barcodes and heterogeneity spacers were trimmed off using tagcleaner v0.16 (72). Trimmed fastq files were loaded into R (v4.0.0) with the DADA2 (73) package version 1.18.0. Quality filtering was performed in DADA2 using the following criteria: *trim=0, maxEE=c(2,2), truncQ=2, rm.phix=TRUE*. Filtered reads were run through the DADA2 Amplicon Sequence Variant (ASV) assignment tool to denoise, cluster and merge the reads. ASVs were assigned a taxonomy from the SILVA version 138.1 rRNA database (74) using the RDP naïve Bayesian classifier (75). The resulting data tables were combined into a phyloseq object using Phyloseq (76).

To remove spurious and likely false-positive ASVs, both an abundance and prevalence filter was applied to the data. ASVs had to contain at least 0.005 % of the total reads to remain in the dataset as well as to be present in at least 1 % of the samples and were otherwise removed. At this step in the pipeline, samples were also removed based on several other criteria such as being a possible sample swaps, >= 8 days in the mail, known duplicates, or poor QC statistics. For this step, samples with less than 4.5K reads OR those which lost more than 50% of reads in the last steps of the DADA2-QC (i.e., those with lots of reads but distributed really in scarce ASV) were removed from the data. Also, samples with 4.5K-6K reads which lost more than 20% of reads in the last steps of the DADA2-QC were excluded. Alpha diversities were calculated based on this filtered phyloseq object. Additionally, a phylogenetic tree was constructed based on the center sequences of each ASV using the phangorn package, and the result was added to the phyloseq object (77). Finally, ASV IDs were recoded to numerical IDs, ordered on ASV abundance within the population.

#### Assessment of general cognition

A neuropsychological assessment battery was introduced in the Rotterdam Study between 2002 and 2005 for evaluating the cognitive function. This battery of tests included the Stroop test (reading, color naming, and interference tasks), a letter-digit substitution task (LDST), a categorical Word Fluency Test (WFT), Purdue Pegboard (PPB) tests for both hands individually and combined, and a 15-word verbal learning test based on Rey’s recall of words (15-WLT). A composite measure of overall cognitive function known as ‘G-factor’ was calculated using principal component analysis, as detailed in previous publications (78). This G-factor consists of scores from the Stroop interference test, LDST, verbal fluency task, PPB test, and 15-WLT delayed recall score.

#### MRI features

MRI scanning has been performed within the Rotterdam Study using a 1.5-T MRI unit equipped with a dedicated eight-channel head coil (Signa HD platform, GE Healthcare, Milwaukee, USA). Brain volumetric measurements, including brain volume, white matter hyperintensity volume, and intracranial volume, were estimated through automated segmentation (79, 80). Left and right hippocampal volumes were obtained using FreeSurfer (version 5.1) and averaged to determine total hippocampal volume. Participants with significant strokes that could potentially affect segmentation were excluded from the MRI marker analysis. Further details regarding MRI scanning and preprocessing can be found elsewhere (81).

### Statistical analysis

#### Association of metabolites with general cognition and MRI markers

To evaluate the association of metabolites with general cognition and MRI markers, we performed linear regression analysis. In the association analyses between general cognition and metabolites, we adjusted the models for age, sex, BMI, and lipid-lowering medication. Among the MRI markers, we selected total brain volume, total hippocampal volume, and total white matter lesions as brain markers of neurodegeneration and vascular health. Natural log transformation and z-transformation (µ = 0, SD = 1) was applied before the linear regression analysis. In the linear models, we adjusted for age at blood collection, the time difference between blood collection and MRI scan, sex, BMI, lipid-lowering medications use, and intracranial volume. We also calculated False Discovery Rate (FDR) by Benjamini Hochberg (82) to determine the statistical significance threshold of associations (FDR < 0.05) between metabolites and general cognition as well as MRI markers. To identify the sex-specific association of metabolites with general cognition and MRI markers, we also performed the regression analysis in sex-stratified samples.

#### Association of metabolites with gut microbial and exposomal features

To perform the association analyses between gut microbiota and circulating metabolites, we performed the central log transformation (CLR) on each of the taxonomic levels of the gut microbiome dataset including phylum, class, order, family, genus, and species using the microbiome package (83). Moreover, we applied z-transformation on the metabolomics profiles (µ=0, SD=1). We performed linear regression analysis to evaluate the association between the transformed plasma levels of metabolites and gut microbial taxa correcting for effects of age, sex, BMI, medication use (proton pump inhibitors, metformin, lipid-lowering medication, and antibiotics), lifestyle factors (smoking, alcohol intake) and technical covariates such as DNA extraction batch, sequencing batch and time of feces in the mail. We also performed linear regression analysis to evaluate the association of individual features included in medication use (31 medications), lifestyle (BMI, alcohol consumption in gram per day, smoking, education level) and clinical factors (diabetes, hypertension, diastolic blood, diastolic blood pressure), where metabolites were used as outcome variable. All these analyses were adjusted for age at blood collection for metabolomics and sex. We applied the significance threshold of 5% FDR in each set of tested features separately.

#### Explained variance of metabolites

In order to calculate the explained variance (EV) of plasma levels of 991 metabolites by genetics, gut microbiota, medication use, lifestyle, and clinical features, we used the Gradient Boosting Decision Tree (GBDT) based machine learning algorithm from LightGBM (V.2.1.2). Bar *et al.* (25) have systematically performed comparison of GBDT and linear based models and reported the high predictive power of GBDT compared to linear models such as LASSO (25). In order to calculate the EV of metabolites based on various features, we therefore adopted the approach described in Bar *et al*. (25). For each group of features, we calculated the EV of each metabolite by using five-fold cross validation. The coefficient of determination (*R^2^*)*100 was interpreted as percentage EV of a metabolite. In the EV calculation for gut microbiota, we used following parameters: learning_rate = 0.005, feature_fraction = 0.2, min_data_in_leaf = 15, metric = l2, early_stopping_rounds = None, n_estimators = 2000, bagging_fraction = 0.8, bagging_freq = 1. To estimate the variance explained by the remaining features (genetics, medication use, lifestyle and clinical features), we used the parameters as predetermined in the LightGBM package: learning_rate = 0.01, max_depth = 5, feature_fraction = 0.8, num_leaves = 25, min_data_in_leaf = 15, metric = L2, early_stopping_rounds = None, n_estimators = 200, bagging_fraction = 0.9, bagging_freq = 5. The genetic, medication, clinical, and lifestyle components were defined as follows: *Genetics:* We used data on single nucleotide polymorphisms (SNPs) for the calculation of percentage EV by genetics. In order to select the SNPs for EV calculation, we adopted a two-step approach. First, we performed a genome-wide association study (GWAS) for all 991 metabolites to prioritize variants with marginal significance of association with metabolites (*P* < 5x10^-8^). GWAS was performed using the HASE software (84). In the summary statistics of the GWAS, we retained variants with imputation quality R^2^ > 0.3 and minor allele frequency (MAF) > 0.05. Furthermore, we performed clumping of SNPs on the summary statistics using the PLINK 1.9 software (85) with a p-value threshold of 5.0x10^-8^ and linkage disequilibrium (LD) threshold (r^2^) of 0.2 in the 500KB region. We observed, in total, 415 independent SNPs reaching a significance p-value threshold of 5.0x10^-8^ for 991 metabolites. We extracted the dosage information from the genotype imputed data for the prioritized SNPs in the RS participants. In the second step, we used the GBDT to calculate the percentage EV of metabolites by genetic features. In calculating the percentage EV of each metabolite, we only used genetic variant features associated with that particular metabolite (*P* <5 x10^-8^) informed by GWAS summary statistics and clumping. We only considered metabolites explained by genetic features with a coefficient of determination (R²) greater than zero and a false discovery rate (FDR) < 0.05 for the p-values of the Spearman correlation coefficient from the GBDT model. In addition, we calculated heritability estimates (*H^2^*) for all 991 metabolites based on the Massively expedited genome-wide heritability analysis (MEGHA) method (86). Due to the small sample size for heritability calculations, we retained heritability estimates of metabolites greater than zero.

*Medication use:* We defined medication intake features based on the intake of medications (Yes/No) information for 31 general medications for which data was recorded in the RS-III cohort at the second follow-up. We included only those medications reported to be used by at least 1 % of our participants (N = 1,068).

*Gut microbiota*: ASV information of all six taxonomic levels including phylum (n = 10), class (n = 17), order (n = 38), family (n = 62), genus (n = 190), and species (n = 151) were used in 922 participants.

*Lifestyle:* In the EV calculation for lifestyle factors, we considered BMI, alcohol consumption in grams per day, smoking (current, former, never), and education level (lower, middle, high). Lifestyle information was available for 1,054 participants with metabolomics data available. *Clinical factors:* Common clinical information including diabetes, hypertension, diastolic blood, diastolic blood pressure was used. Full information on clinical parameters was available for 1,054 participants with metabolomics data.

## Data Availability

Due to ethical and legal restrictions, individual‐level data from the Rotterdam Study (RS) are only available upon request to the data manager of the Rotterdam Study Frank van Rooij and subject to local rules and regulations. This includes submitting a proposal to the management team of RS, where upon approval, analysis needs to be done on a local server with protected access, complying with GDPR regulations. All other data produced in the present study are contained in the manuscript or are available upon reasonable request to the authors.

## ACKNOWLEDGEMENTS

The Rotterdam Study is supported by the Erasmus MC University Medical Center and Erasmus University Rotterdam, the Netherlands Organization for Scientific Research (NWO), the Netherlands Organization for Health Research and Development (ZonMW), the Research Institute for Diseases in the Elderly (RIDE), the Ministry of Education, Culture and Science, the Ministry of Health, Welfare and Sport, The European Commission (DGXII), the Netherlands Genomics Initiative (NGI), and the Municipality of Rotterdam.

This project was enabled in part by the Alzheimer’s Gut Microbiome Project (AGMP), supported by the National Institute on Aging grants: 1U19AG063744 and 3U19AG063744-04S1, awarded to Dr. Kaddurah-Daouk at Duke University in partnership with multiple academic institutions. As such, the investigators within the AGMP not listed in this publication’s authors’ list, provided analysis-ready data, but did not participate in designing the study, conducting the analyses or writing of this manuscript. A listing of AGMP Investigators can be found at https://alzheimergut.org/meet-the-team/.

Metabolomics data used in preparation of this article were generated by the Alzheimer’s Disease Metabolomics Consortium (ADMC). As such, the investigators within the ADMC other than named authors provided data but did not participate in analysis or writing of this report. A complete listing of ADMC investigators can be found at: https://sites.duke.edu/adnimetab/team/. The NIA supported the Alzheimer’s Disease Metabolomics Consortium which is a part of NIA’s national initiatives AMP-AD and M^2^OVE-AD (R01 AG046171, RF1AG059093, RF1AG058942, RF1 AG051550, 3U01AG061359, 3U01 AG024904-09S4). In addition, this study used data, of which the production was funded by ZonMW Memorabel (project number #733050814).

