## Supplementary Material and Figures for "The blood metabolome of cognitive function and brain health in middle-aged adults – influences of genes, gut microbiome, and exposome"

### Details of metabolomics data acquisition at Metabolon Inc.

***Sample Accessioning:*** Following receipt, samples were immediately stored at -80$^{o}$C. Each sample received was accessioned into the Metabolon Laboratory Information Management System (LIMS) and was assigned a unique identifier. This identifier was used to track all sample handling, tasks, results, etc. The samples (and all derived aliquots) were tracked by the LIMS system.

***Sample preparation:*** Samples were prepared using the automated MicroLab STAR® system from Hamilton. Several recovery standards were added prior to the first step in the extraction process for quality control (QC) purposes. To remove protein, dissociate small molecules bound to protein or trapped in the precipitated protein matrix, and to recover chemically diverse metabolites, proteins were precipitated with methanol under vigorous shaking for 2 min (Glen Mills GenoGrinder 2000) followed by centrifugation. The resulting extract was divided into five fractions: two for analysis by two separate reverse phase (RP)/UPLC-MS/MS methods with positive ion mode electrospray ionization (ESI), one for analysis by RP/UPLC-MS/MS with negative ion mode ESI, one for analysis by HILIC/UPLC-MS/MS with negative ion mode ESI, and one sample was reserved for backup. Samples were placed briefly on a TurboVap® (Zymark) to remove the organic solvent. The sample extracts were stored overnight under nitrogen before preparation for analysis.

***Quality assurance/quality control:*** Several types of controls were analyzed in concert with the experimental samples: a pooled matrix sample of well-characterized human plasma served as a technical replicate throughout the data set; extracted water samples served as process blanks; and a cocktail of QC standards that were carefully chosen not to interfere with the measurement of endogenous compounds were spiked into every analyzed sample, allowed instrument performance monitoring and aided chromatographic alignment. Instrument variability was determined by calculating the median relative standard deviation (RSD) for these standards. Overall process variability was determined by calculating the median RSD for all endogenous metabolites (i.e., non-instrument standards) present in 100% of the pooled matrix samples. Experimental samples were randomized across the platform run with QC samples spaced evenly among the injections.

***Ultrahigh performance liquid chromatography-tandem mass spectroscopy (UPLC-MS/MS):*** All methods utilized a Waters ACQUITY ultra-performance liquid chromatography (UPLC) and a Thermo Scientific Q-Exactive high resolution/accurate mass spectrometer interfaced with a heated electrospray ionization (HESI-II) source and Orbitrap mass analyzer operated at 35,000 mass resolution. The sample extracts were dried then reconstituted in solvents compatible to each of the four methods. Each reconstitution solvent contained a series of standards at fixed concentrations to ensure injection and chromatographic consistency. One aliquot was analyzed using acidic positive ion conditions, chromatographically optimized for more hydrophilic compounds. In this method, the extract was gradient eluted from a C18 column (Waters UPLC BEH C18-2.1x100 mm, 1.7 µm) using water and methanol, containing 0.05% perfluoropentanoic acid (PFPA) and 0.1% formic acid (FA). Another aliquot was also analyzed using acidic positive ion conditions; however, it was chromatographically optimized for more hydrophobic compounds. In this method, the extract was gradient eluted from the same afore mentioned C18 column using methanol, acetonitrile, water, 0.05% PFPA and 0.01% FA and was operated at an overall higher organic content. Another aliquot was analyzed using basic negative ion optimized conditions using a separate dedicated C18 column. The basic extracts were gradient eluted from the column using methanol and water, however with 6.5mM Ammonium Bicarbonate at pH 8. The fourth aliquot was analyzed via negative ionization following elution from a HILIC column (Waters UPLC BEH Amide 2.1x150 mm, 1.7 µm) using a gradient consisting of water and acetonitrile with 10mM Ammonium Formate, pH 10.8. The MS analysis alternated between MS and data-dependent $MS^n$ scans using dynamic exclusion. The scan range varied slightly between methods but covered 70-1000 m/z. Raw data files are archived and extracted as described below.

***Bioinformatics:*** The informatics system consisted of four major components, the LIMS, the data extraction and peak-identification software, data processing tools for QC and compound identification, and a collection of information interpretation and visualization tools for use by data analysts. The hardware and software foundations for these informatics components were the LAN backbone, and a database server running Oracle 10.2.0.1 Enterprise Edition.

***Data extraction and compound identification:*** Raw data was extracted, peak-identified and QC processed using Metabolon's hardware and software. Compounds were identified by comparison to library entries of purified standards or recurrent unknown entities. Metabolon maintains a library based on authenticated standards that contains the retention time/index (RI), mass to charge ratio (m/z), and chromatographic data (including MS/MS spectral data) on all molecules present in the library. Furthermore, biochemical identifications are based on three criteria: retention index within a narrow RI window of the proposed identification, accurate mass match to the library +/- 10 ppm, and the MS/MS forward and reverse scores between the experimental data and authentic standards. The MS/MS scores are based on a comparison of the ions present in the experimental spectrum to the ions present in the library spectrum. While there may be similarities between these molecules based on one of these factors, the use of all three data points can be utilized to distinguish and differentiate biochemicals. More than 3300 commercially available purified standard compounds have been acquired and registered into LIMS for analysis on all platforms for determination of their analytical characteristics. Additional mass spectral entries have been created for structurally unnamed biochemicals, which have been identified by virtue of their recurrent nature (both chromatographic and mass spectral). These compounds have the potential to be identified by future acquisition of a matching purified standard or by classical structural analysis.

***Curation:*** A variety of curation procedures were carried out to ensure accurate and consistent identification of true chemical entities, and to remove those representing system artifacts, mis-assignments, and background noise. Metabolon data analysts use proprietary visualization and interpretation software to confirm the consistency of peak identification among the various samples. Library matches for each compound were checked for each sample and corrected if necessary.

***Metabolite quantification and data normalization:*** Peaks were quantified using area-under-the-curve. For studies spanning multiple days, a data normalization step was performed to correct variation resulting from instrument inter-day tuning differences. Essentially, each compound was corrected in run-day blocks by registering the medians to equal one (1.00) and normalizing each data point proportionately (termed the "block correction").

**Additional details of preprocessing data from gut microbiome profiling**

The 16S rRNA sequencing data from the samples of the RS-III cohort have been processed as follows.

Barcodes were separated from the reads and both ends were pasted together. These barcodes were then used to separate (demultiplex) the reads for each sample using an in-house developed script, allowing 1 error in each 12 nucleotide half of the 24 nucleotides long barcode. Reads were then cleaned of primer sequences and heterogeneity spacers using tag cleaner version 0.16 (1). Trimmed reads were then imported into the DADA2 R package version 1.18.0 (2). Samples without reads were removed and remaining reads were used as input for the filter step of DADA2. This filtering step removed reads with an expected error rate higher than 2 in both the forward and reverse reads, as well as any reads which had at least one or more ambiguous bases in them ("N"). Additionally, reads were truncated if a low-quality base was encountered (Q-score =< 2). Next, reads were denoised by clustering them together based on similarity, starting with the most abundant read. All other reads with similarity within 10% were aligned against this cluster and included if the abundance p-value (likelihood of the read being too abundant to be explained by predicted errors in the main cluster sequence) was below the default threshold. The algorithm was then repeated with the remaining reads for the next abundant cluster, and so forth. Due to the nature of the algorithm requiring multiples of the same read, singletons are automatically excluded. The forward and reverse of the denoised reads were then merged if the overlap between them was a 100% match. Reads which were truncated in the filter step of DADA2 were also more likely to get removed in this step, due to possibly not having an overlap anymore. Next, the remaining chimeric sequences were removed using remove BimeraDenovo in consensus mode within DADA2. These cleaned, clustered reads (hereafter named Amplicon Sequence Variant, ASV) were then used as input for the RDP naive Bayesian classifier, which was trained on the SILVA version 138.1 microbial database (3, 4). Taxonomy for Kingdom through Genus was assigned if the bootstrap confidence score was above 50, meaning that a random part of the read could be assigned to those taxa at least 50% of the time. Species assignment was only reported if the ASV could be matched to a species for 100% and could not be mapped to other species either; otherwise “NA” was reported after the genus name. The ASV table, taxonomy table, and metadata were then combined into a phyloseq object for ease of analysis (5).

As described in **Methods**, further steps were taken to remove spurious and likely false-positive ASVs, filter samples, and for phylogenetic tree construction, which was added to the phyloseq object.

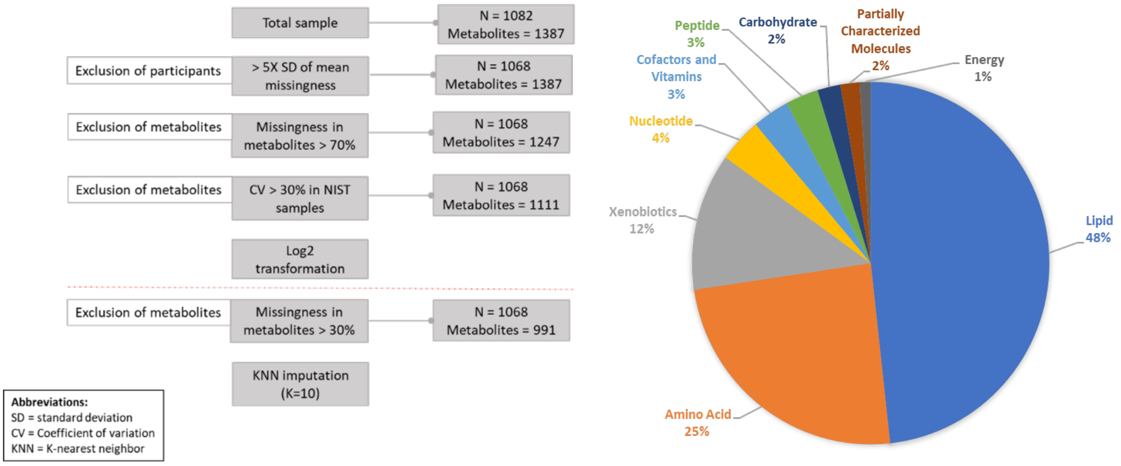


**Supplementary Figure 1:** Preprocessing of the metabolomics data and metabolite classes. (A) flowchart showing the steps performed during preprocessing of the metabolomics dataset. (B) Pi-chart showing the percentage of different metabolite classes covered in the metabolomics data set.

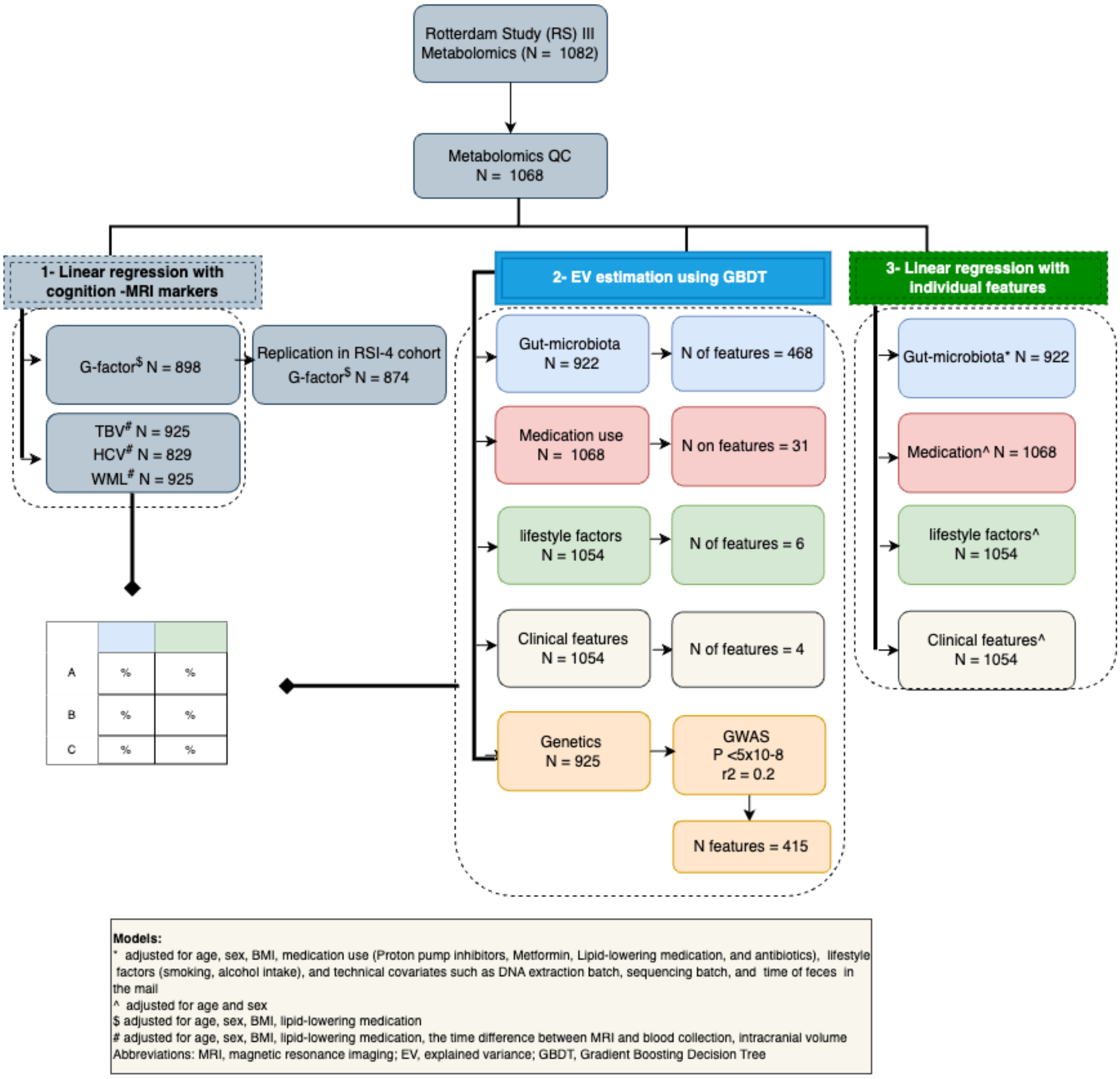


**Supplementary Figure 2:** Detailed flowchart of study design and analysis steps.

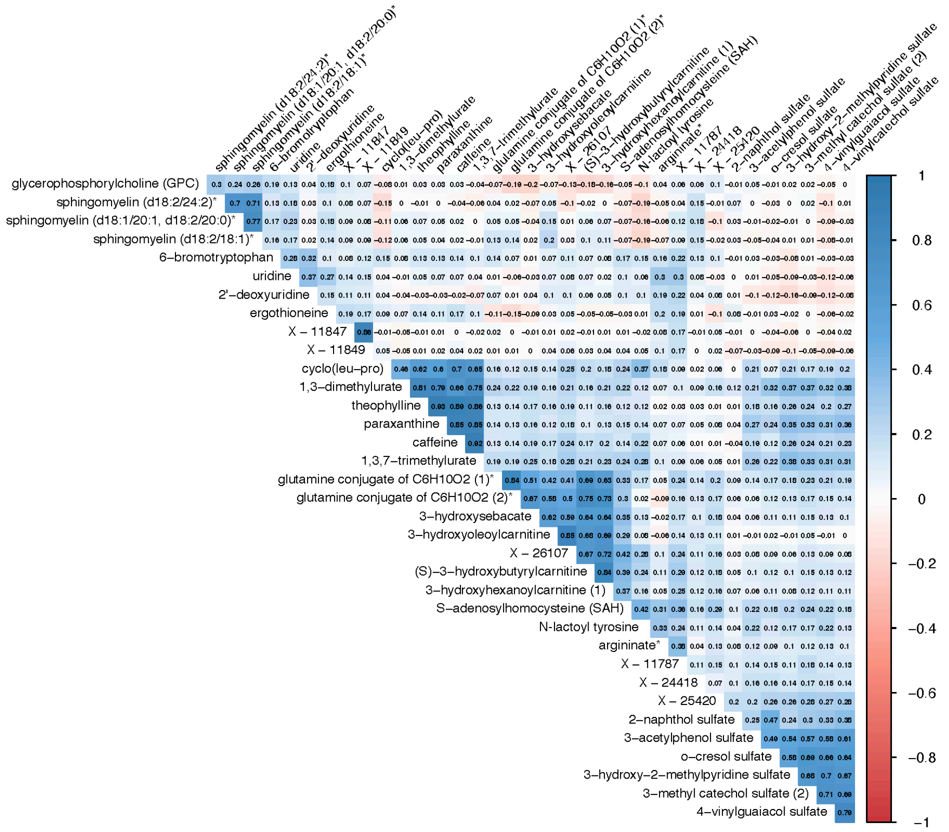


**Supplementary Figure 3:** Spearman correlation matrix of metabolites associated with general cognition and MRI markers. Note: N-lactoyltyrosine is the updated annotation of a metabolite that was annotated as 1-carboxyethyltyrosine in the original data set. This correction was provided by Metabolon.

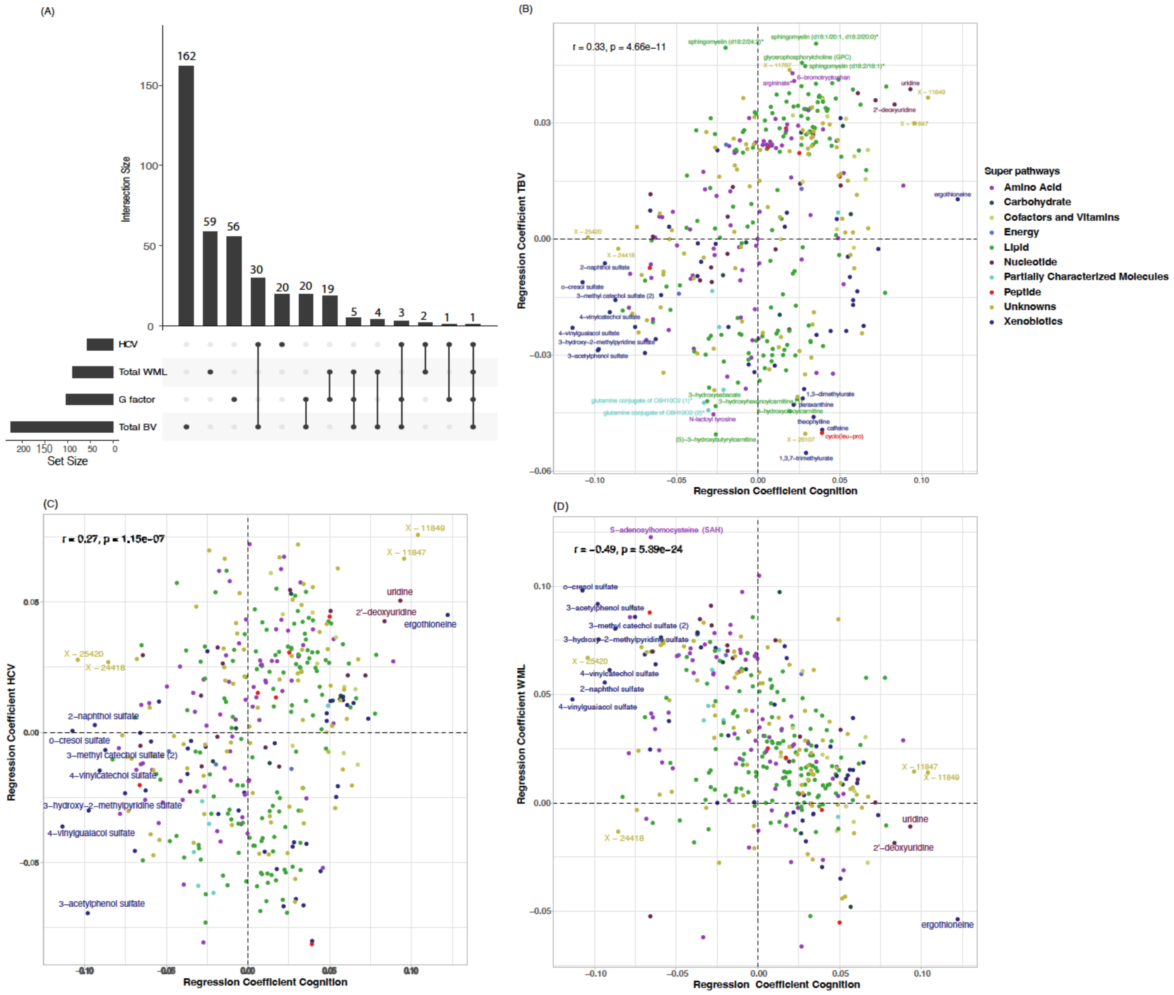


**Supplementary Figure 4:** Concordance of metabolite signatures between cognition and MRI phenotypes. Correlation plots show the regression coefficients of the metabolites that are associated with general cognition, total brain volume, total hippocampal volume, and total white matter lesions at $P < 0.05$ (nominal significance). The color indicates the class of metabolites. Abbreviations: BV – brain volume; HCV – hippocampal volume; WML – white matter lesions; G-factor – general cognition.

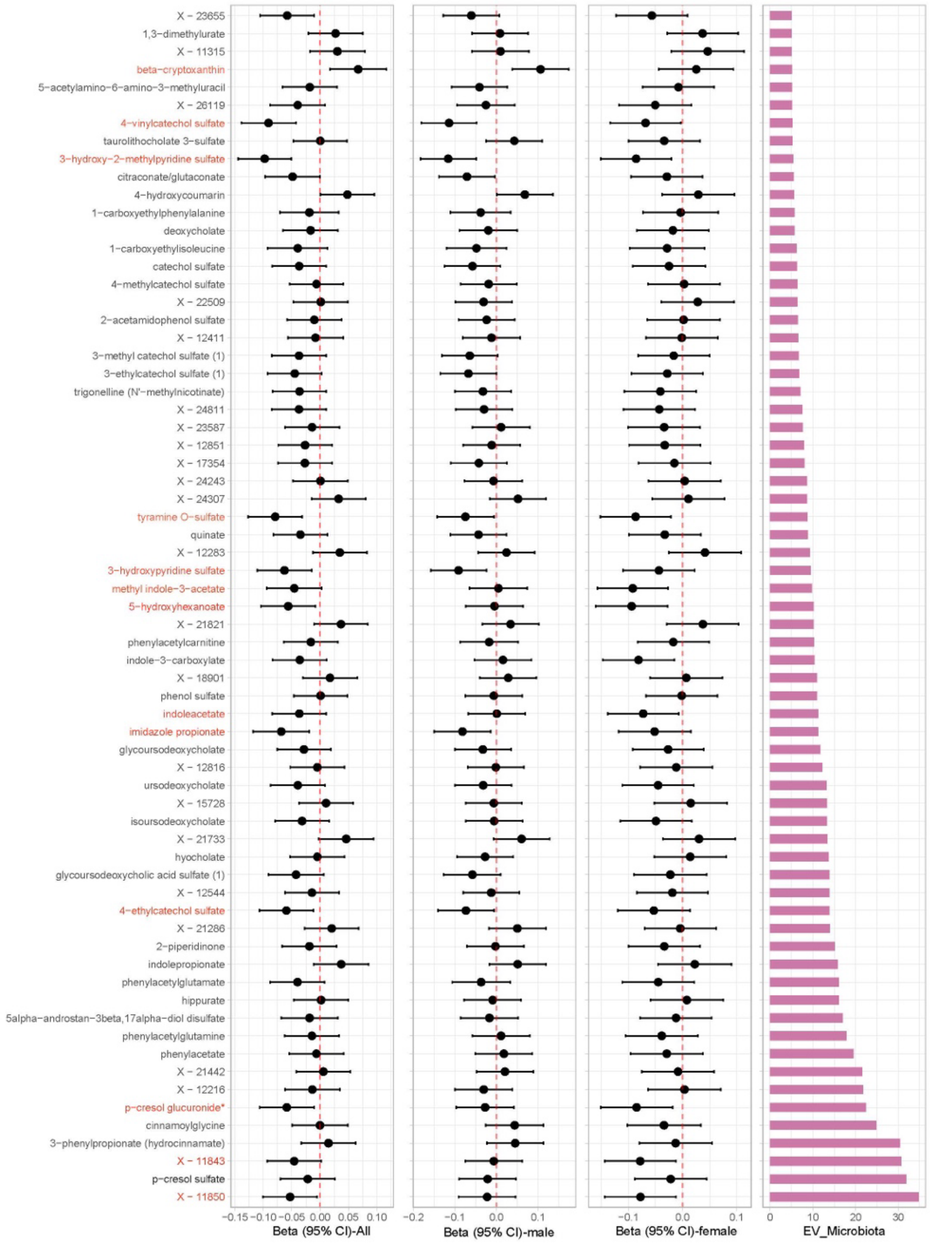


**Supplementary Figure 5:** Relationship of metabolites strongly influenced by the gut microbiome (EV $\geq$ 5%) with metabolites nominally associated ($P$ < 0.05) with general cognition (shown in red). Note: The original annotation for 1-carboxyethylphenylalanine and 1-carboxyethylisoleucine has been corrected to N-lactoylphenylalanine and N-lactoylisoleucine meanwhile.

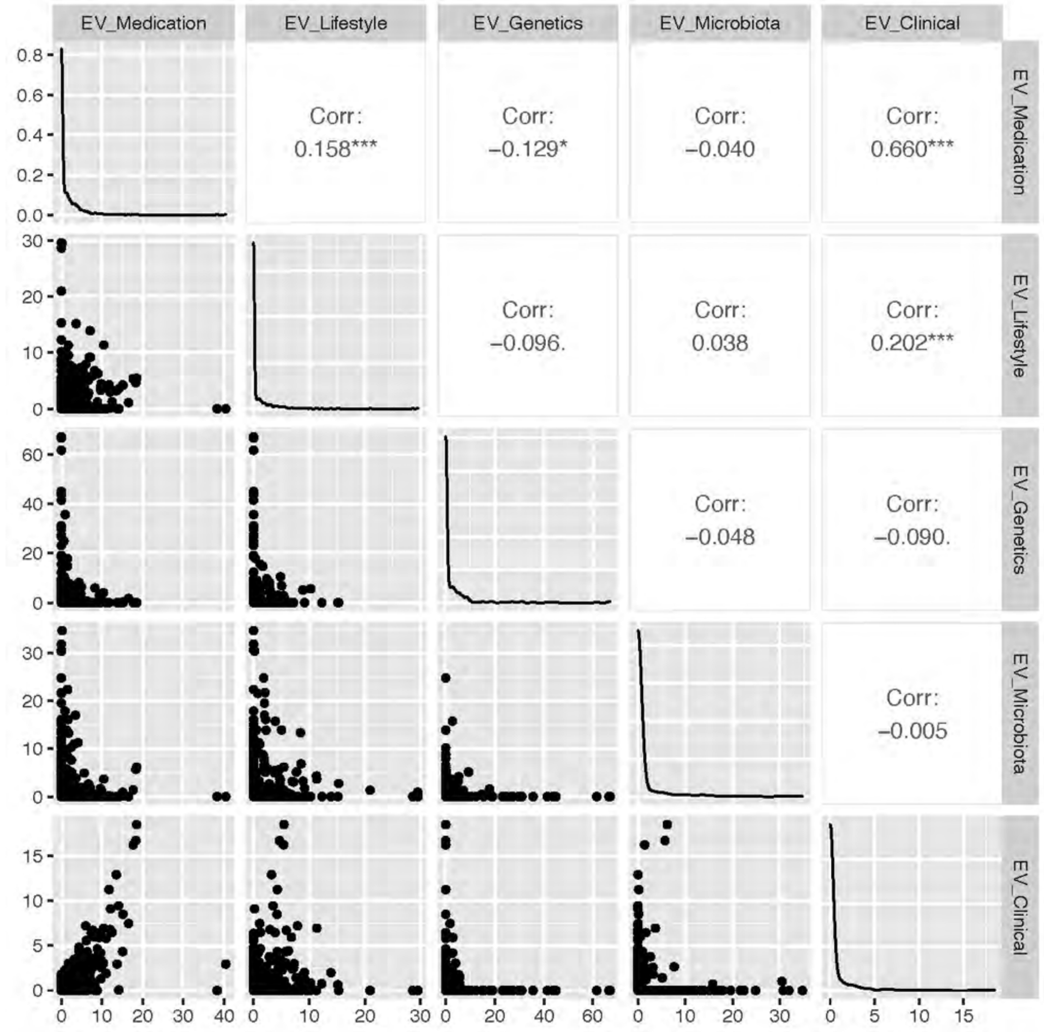


**Supplementary Figure 6:** Correlation plot between explained variance (EV) by different tested features. For the correlation analysis, we only considered metabolites EV > 0 and false discovery rate (FDR) of spearman correlation < 0.05 for every tested class of features.

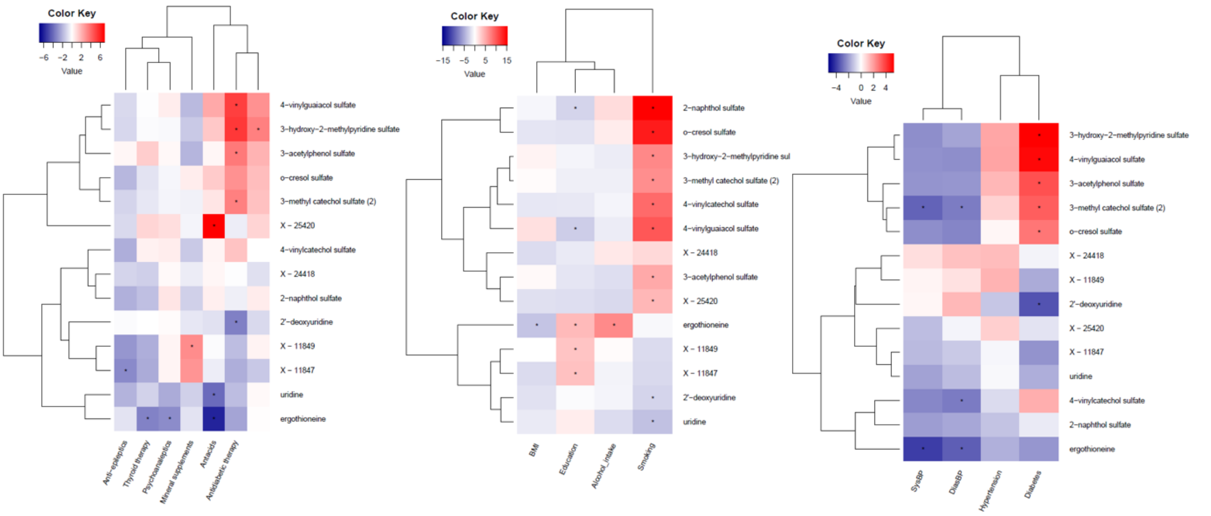


**Supplementary Figure 7:** Relationship of cognition-associated metabolites with medication use (A), lifestyle (B), and clinical factors (C). The color key indicates the direction of the association. A star in the heatmap represents a significant association (FDR < 0.05) based on the association between all tested metabolites with each set of tested features.

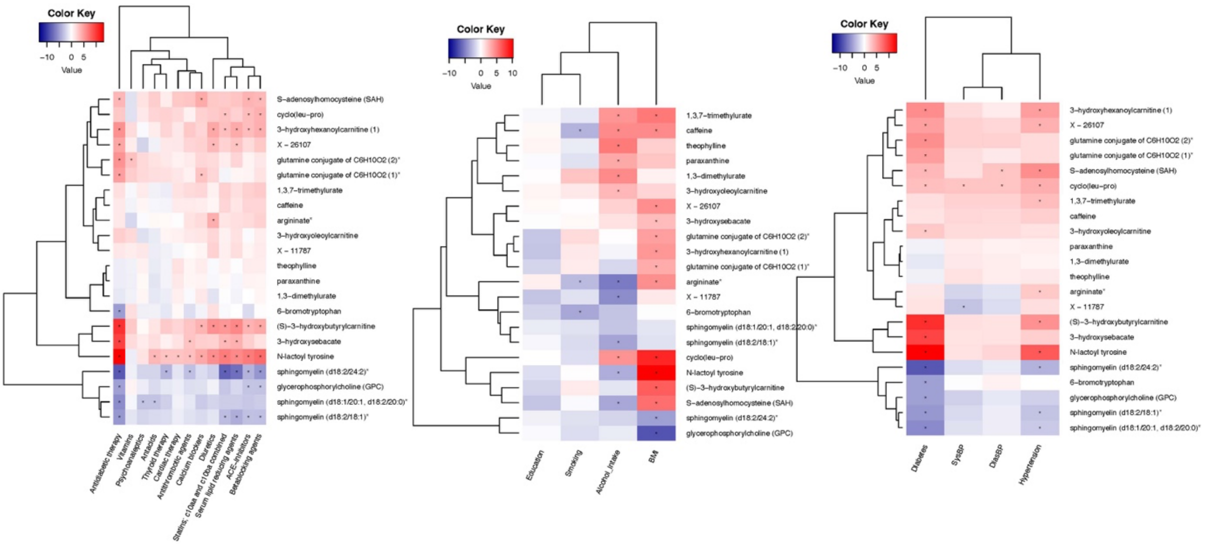


**Supplementary Figure 8:** Relationship of imaging marker-associated metabolites with medication use (A), lifestyle (B), and clinical factors (C). The color indicates the direction of the association. A star in the heatmap represents a significant association (FDR < 0.05) based on the association between all tested metabolites with each set of tested features. Note: N-lactoyltyrosine is the updated annotation of a metabolite that was annotated as 1-carboxyethyltyrosine in the original data set. This correction was provided by Metabolon.

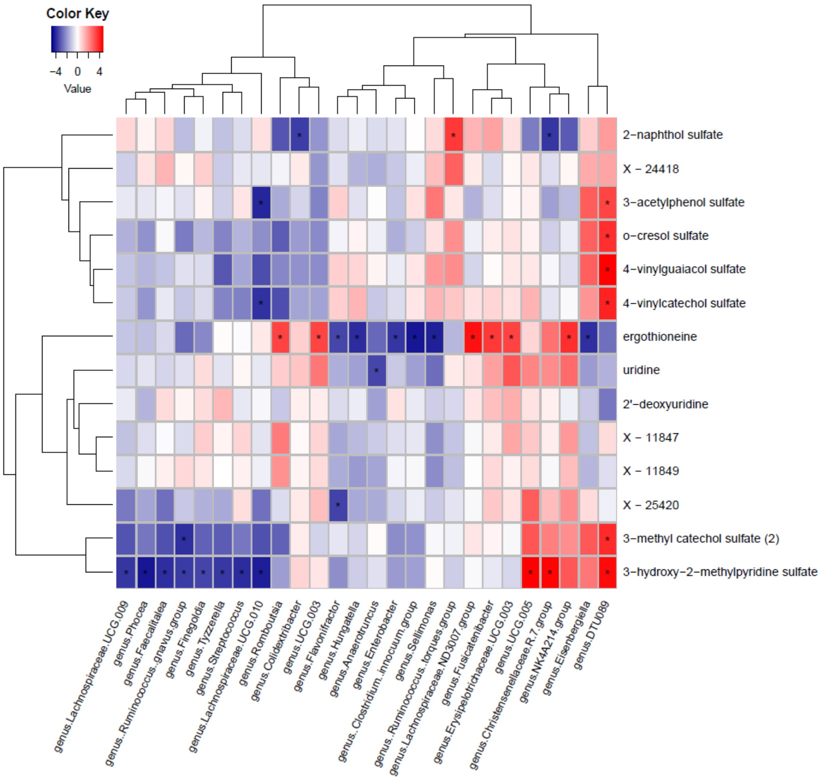


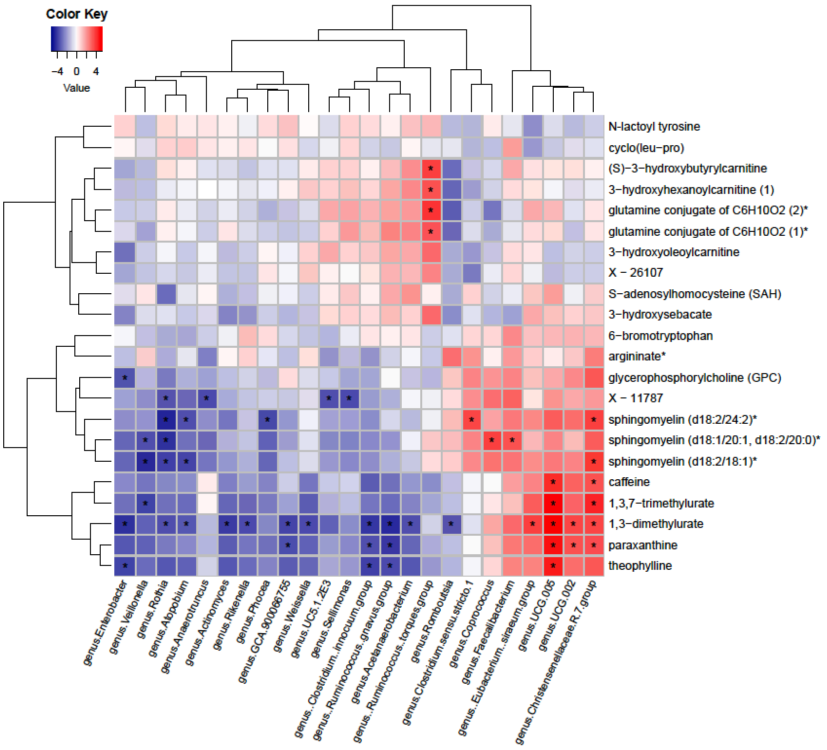


**Supplementary Figure 9:** Relationship of general cognition and brain imaging associated metabolites with gut microbiota. (A) Association between metabolites related to general cognition with gut microbiota at the genus level. (B) Association between metabolites associated with imaging markers with gut microbiota at the genus level. A star in the heatmap represents a significant association (FDR < 0.05) based on the association between all tested metabolites with all microbial taxonomic levels. Note: N-lactoyltyrosine is the updated annotation of a metabolite that was annotated as 1-carboxyethyltyrosine in the original data set. This correction was provided by Metabolon.

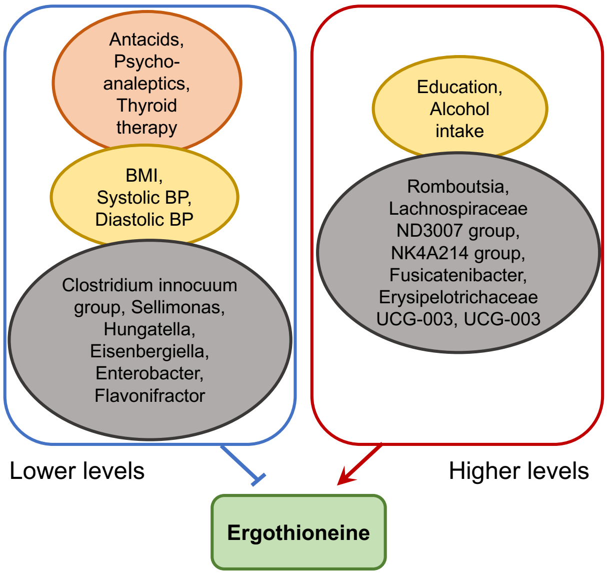


**Supplementary Figure 10:** Overview of significant associations (FDR < 0.05) of ergothioneine blood levels with gut microbial and exposomal factors in this study.

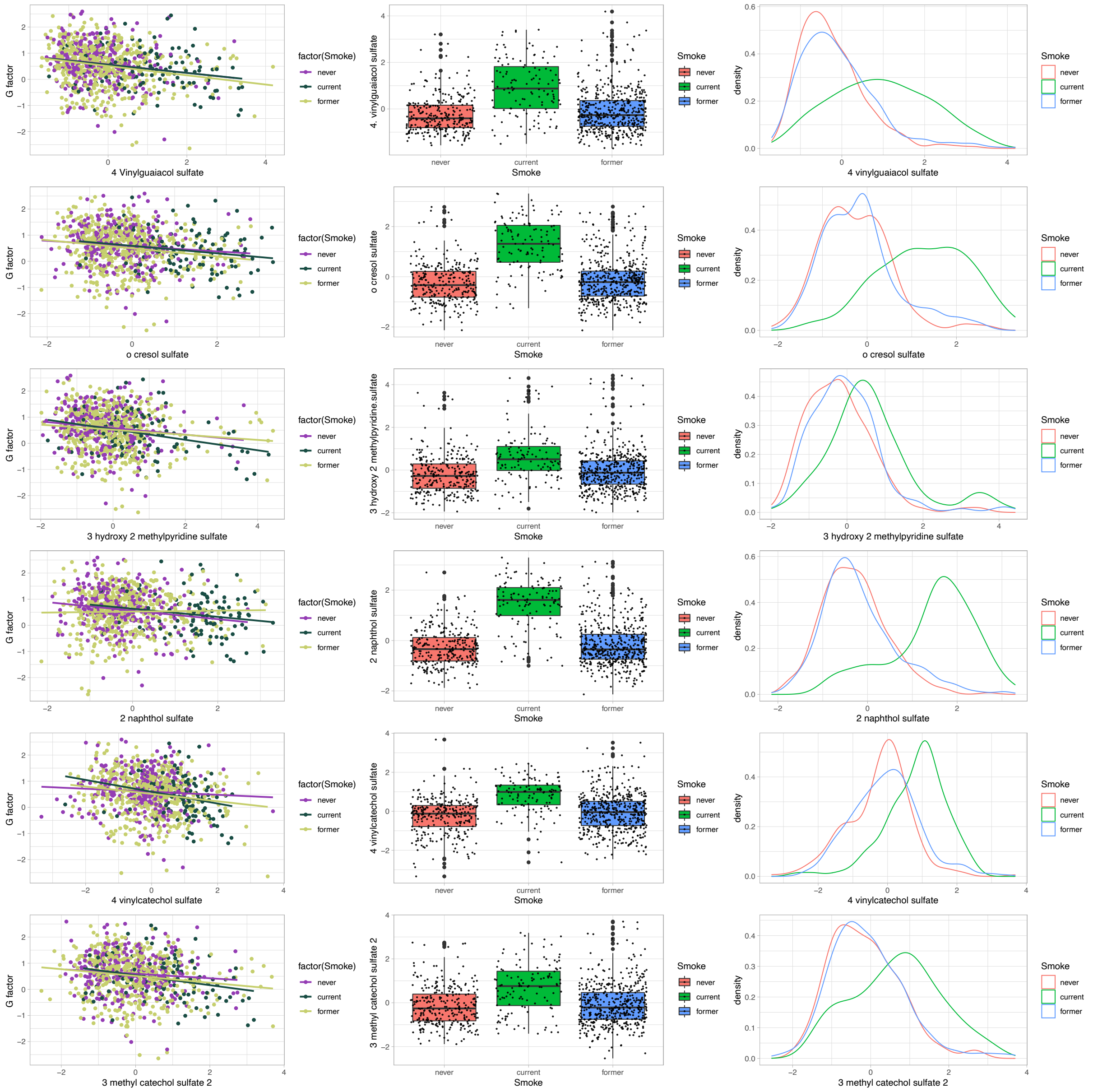


**Supplementary Figure 11:** Distribution of sulfated xenobiotic metabolites (4-vinylguaiacol sulfate, o-cresol sulfate, 3-hydroxy-2-methylpyridine sulfate, 2-naphthol sulfate, 4-vinylcatechol sulfate, 3-methylcatechol sulfate) associated with general cognition and smoking.
